# Characterizing the blood stage antimalarial activity of tafenoquine in healthy volunteers experimentally infected with *Plasmodium falciparum*

**DOI:** 10.1101/2022.11.21.22282610

**Authors:** Bridget E. Barber, Azrin N. Abd-Rahman, Rebecca Webster, Adam J. Potter, Stacey Llewellyn, Louise Marquart, Nischal Sahai, Indika Leelasena, Geoffrey W. Birrell, Michael D. Edstein, G. Dennis Shanks, David Wesche, Joerg J. Moehrle, James S. McCarthy

## Abstract

**Background:** The long acting 8-aminoquinoline tafenoquine may be a good candidate for mass drug administration if it exhibits sufficient blood stage antimalarial activity at doses low enough to be tolerated by glucose 6-phosphate dehydrogenase (G6PD) deficient individuals.

**Methods:** Healthy G6PD-normal adults were inoculated with *Plasmodium falciparum* 3D7-infected erythrocytes on day 0. Different single oral doses of tafenoquine were administered on day 8. Parasitemia, and concentrations of tafenoquine and the 5,6-orthoquinone metabolite in plasma/whole blood/urine were measured and standard safety assessments performed. Curative artemether-lumefantrine therapy was administered if parasite regrowth occurred, or on day 48±2. Outcomes were parasite clearance kinetics, pharmacokinetic and pharmacokinetic/pharmacodynamic (PK/PD) parameters from modelling, and dose simulations in a theoretical endemic population.

**Results:** Twelve participants were inoculated and administered 200 mg (n=3), 300 mg (n=4), 400 mg (n=2), or 600 mg (n=3) tafenoquine. The parasite clearance half-life with 400 mg or 600 mg (5.4 h and 4.2 h respectively) was faster than with 200 mg or 300 mg (11.8 h and 9.6 h respectively). Parasite regrowth occurred after dosing with 200 mg (3/3 participants) and 300 mg (3/4 participants), but not after 400 mg or 600 mg. Simulations using the PK/PD model predicted that 460 mg and 540 mg would clear parasitaemia by a factor of 10^6^ and 10^9^, respectively, in a 60 kg adult.

**Conclusions:** Although a single dose or tafenoquine exhibits potent *P. falciparum* blood stage antimalarial activity, the estimated doses to effectively clear asexual parasitemia will require prior screening to exclude G6PD deficiency.

**Main point:** A single oral dose of tafenoquine is effective against blood stage *Plasmodium falciparum* infection. However, as the estimated dose to clear asexual parasitaemia is ≥460 mg (in adults), prior screening for glucose 6-phosphate dehydrogenase deficiency will be required.

## INTRODUCTION

The global burden of malaria remains high, with an estimated 241 million cases and 627,000 deaths in 2020 [1]. The emergence of artemisinin-resistant *Plasmodium falciparum* in Southeast Asia [2], and more recently in East Africa [3], threatens the utility of the current first line artemisinin combination therapies and progress toward malaria eradication. There is growing recognition for the need to treat both asymptomatic and symptomatic malaria infections to reduce the parasite reservoir [4] and to slow the spread of drug-resistant parasites [5]. Mass drug administration (MDA) in endemic populations is one approach to rapidly reduce malaria morbidity and mortality and interrupt transmission. The World Health Organization (WHO) defines malaria MDA as the administration of a full dose of antimalarial treatment to an entire population in a given area, except those in whom the medicine is contraindicated [6]. Further, medicines used for MDA should be of proven efficacy in the implementation area, have a long half-life, and be different to those medicines used for first line treatment [6].

The 8-aminoquinoline tafenoquine is a long acting analogue of primaquine that is currently approved for malaria chemoprophylaxis and for radical cure of vivax malaria [7]. The fact that tafenoquine has a very long elimination half-life (approximately 16 days [8]) and broad activity against all parasite lifecycle stages [9], indicates that it may be a suitable candidate for use in MDA. However, although the blood schizonticidal activity of tafenoquine has been established in humans [10, 11], the dose required to clear asexual *P. falciparum* parasites has not yet been determined. Furthermore, as with primaquine, the use of tafenoquine is limited by the risk of hemolysis in glucose-6-phosphate dehydrogenase (G6PD) deficient individuals, and administration of tafenoquine at currently recommended doses (200 mg for chemoprophylaxis and 300 mg for *P. vivax* radical cure) requires prior G6PD testing, which can be costly and logistically difficult in low- and middle-income countries. However, the fact that the hemolytic potential of tafenoquine appears to be dose-dependent [12] suggests that low dose tafenoquine may be safe in G6PD-deficient individuals, in a similar manner to low dose primaquine [13].

We hypothesized that if a sufficiently low effective dose of tafenoquine was identified, this might allow tafenoquine to be used for MDA in endemic populations, without the need for G6PD testing prior to administration. The purpose of the current study was to use the induced blood stage malaria (IBSM) model to determine the minimum single oral dose of tafenoquine required to effectively clear asexual blood stage *P. falciparum*.

## METHODS

### Study design and participants

This was an open label, randomized, clinical trial using the IBSM model. Healthy malaria naïve males and females (non-pregnant, non-lactating) aged 18-55 years were eligible for inclusion (Text S2). All participants were required to have normal G6PD activity at screening (normal range 7.0-20.5 U/g hemoglobin). The study was conducted at the University of the Sunshine Coast Clinical Trials Unit (Morayfield, Australia) and was registered on the Australian and New Zealand Clinical Trials Registry (ACTRN12620000995976).

This study was approved by the QIMR Berghofer Medical Research Institute Human Research Ethics Committee (P3646) with mutual recognition by the Australian Departments of Defence and Veterans’ Affairs Human Research Ethics Committee (194-19). All participants gave written informed consent before enrollment.

### Procedures

Participants were inoculated intravenously with approximately 2800 viable *P. falciparum* 3D7 infected erythrocytes on day 0 [14]. Parasitemia was monitored throughout the study by quantitative PCR (qPCR) targeting the gene encoding *P. falciparum* 18S rRNA [15]. A single oral dose of tafenoquine (Kodatef^®^, Biocelect) was administered on day 8. All participants in cohort 1 received the same tafenoquine dose (300 mg), with no randomization performed. Participants in cohorts 2 and 3 were randomized to a dose group on day 8. The randomization schedule was generated using STATA 15. No blinding was performed.

Plasma, venous and capillary whole blood, and urine concentrations of tafenoquine (and the 5, 6-orthoquinone metabolite) were measured by liquid chromatography tandem mass spectrometry [16]. Participants received a curative course of artemether-lumefantrine (Riamet^®^, Novartis Pharmaceuticals) if parasite regrowth was detected or on day 48±2. Safety assessments, including monitoring of adverse events (AEs), vital signs, hematology and biochemistry, physical examination, and electrocardiographs, were performed throughout the study.

### Outcomes and Statistical analysis

Analysis of parasite clearance kinetics following tafenoquine was performed in R 3.5.0 and RStudio 1.1.447. The parasite reduction ratio over a 48 h period (PRR_48_) and parasite clearance half-life (PCt_1/2_) were estimated using the slope of the optimal fit for the log-linear relationship of the parasitemia decay [17]. Non-compartmental pharmacokinetic analysis was performed using PKanalix 2019R1 (Lixoft).

Pharmacokinetic (PK) and pharmacokinetic/pharmacodynamic (PK/PD) modelling, and simulations were conducted within R 3.6.3 combined to the IQRtools package 1.7.2 and MonolixSuite 2019R1 (Text S1). PK/PD analyses were performed using non-linear mixed effects models. A population PK model was developed to obtain individual PK parameter estimates that adequately described the observed individual PK profiles. The PK/PD model was then built using the individual PK parameter estimates as regression parameters. Key efficacy parameters including the minimum inhibitory concentration (MIC) and the minimal parasiticidal concentration that achieved 90% of the maximum effect (MPC_90_) were derived from the final PK/PD model.

Simulations were performed in a theoretical endemic population to predict the dose of tafenoquine that would clear parasitemia by a factor of 10^6^ and 10^9^ in all patients, and achieve adequate parasitological response on day 42 (APR_42_) with 90% probability (Text S1). Population parameters were sampled from the uncertainty distribution, and individual parameters were sampled from the inter-individual variability (variability of E_max_ was assumed to be 20%). Single doses of 1 to 40 mg/kg tafenoquine (with an increment of 1 mg/kg) were simulated in 20 trials of 100 patients each with body weight of 5, 10, 15, 20, 30, 40, 50 and 60 kg. Baseline parasitemia was assumed to be log-normally distributed around a median of 10^7^ parasites/mL, with inter-individual variability of 60% [18]. The PK and PK/PD relationship were assumed to be the same in patients as in the volunteers, and parasites were assumed to grow exponentially in patients at a growth rate of 0.048 h^-1^ (equivalent to a multiplication rate of approximately 10-fold per asexual cycle of 48 hours) [19, 20].

## RESULTS

### Participants

The study was conducted from 27 October 2020 to 09 April 2021. Twelve participants were enrolled across three cohorts (Figure 1). Participants were healthy malaria-naïve males (n=6) or females (n=6) aged 19 to 53 years (Table 1). All 3 participants in cohort 1 received 300 mg tafenoquine while participants in cohorts 2 and 3 were randomized to a dose group (200 mg, n=3; 300 mg, n=1; 400 mg, n= 2; or 600 mg, n=3). Doses were selected following review of interim data to optimally characterize the PK/PD relationship. All participants received the allocated treatment, completed the study and were included in the analysis of study endpoints.

**Table 1.** Demographic profile of participants.

|  |  | Tafenoquine 200 mg | Tafenoquine 300 mg | Tafenoquine 400 mg | Tafenoquine 600 mg | Total |
| --- | --- | --- | --- | --- | --- | --- |
|  |  | (N=3) | (N=4) | (N=2) | (N=3) | (N=12) |
| Sex [n (%)] | Female | 1 (33) | 3 (75) | 2 (100) | 0 (0) | 6 (50) |
|  | Male | 2 (67) | 1 (25) | 0 (0) | 3 (100) | 6 (50) |
| Race [n (%)] | White | 2 (67) | 4 (100) | 2 (100) | 3 (100) | 11 (92) |
|  | Asian | 1 (33) | 0 (0) | 0 (0) | 0 (0) | 1 (8) |
| Age [years] | Median (range) | 40 (28 - 53) | 34 (20 - 45) | 31 (20 - 41) | 23 (19 - 23) | 30 (19 - 53) |
| Body weight [kg] | Median (range) | 87 (61 - 95) | 81 (71 - 108) | 52 (51 - 53) | 67 (65 - 100) | 72 (51 - 108) |
| Body mass index [kg/m<sup>2</sup>] | Median (range) | 29 (26 - 30) | 27 (27 - 31) | 20 (20 - 21) | 21 (20 - 30) | 27 (20 - 31) |

**Figure 1.**
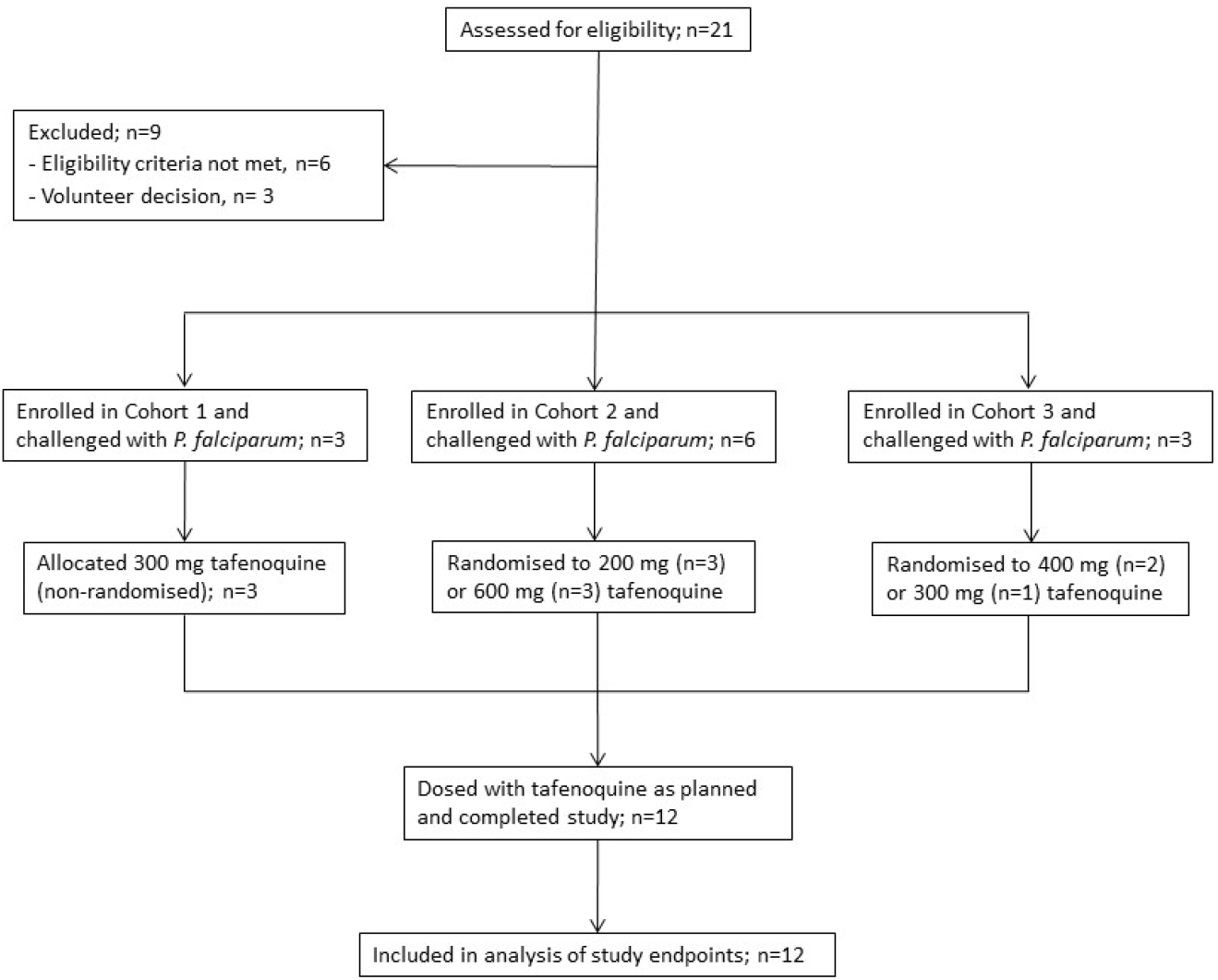
Trial profile. Eligible participants were enrolled in one of three cohorts. All participants in cohort 1 were allocated the same dose of tafenoquine (300 mg). Participants in cohorts 2 and 3 were randomized within each cohort to a dose group on the day of dosing with tafenoquine (8 days following challenge with blood-stage *P. falciparum*). All participants completed the study and were included in the analysis of study endpoints.

### Parasite clearance kinetics and regrowth

The progression of parasitemia following intravenous inoculation was consistent between participants up to day 8, when a single oral dose of tafenoquine was administered (Figure 2). An initial reduction in parasitemia occurred in all participants following tafenoquine, with a trend of an increased rate of parasite clearance with increasing dose of tafenoquine (Table 2). Parasite regrowth occurred after dosing with 200 mg (3/3 participants; 8-22 days post-dose) and 300 mg (3/4 participants; 18-27 days post-dose) tafenoquine but not after dosing with 400 mg or 600 mg tafenoquine up to day 48 when definitive antimalarial treatment with artemether-lumefantrine was initiated (Figure 2).

**Table 2.** Parasite clearance parameters following single dose tafenoquine administration to healthy volunteers with induced *P. faiciparum* parasitemia.

|  | Tafenoquine 200 mg<br>(N=3<sup>a</sup>) | Tafenoquine 300 mg<br>(N=4) | Tafenoquine 400 mg<br>(N=2) | Tafenoquine 600 mg<br>(N=3) |
| --- | --- | --- | --- | --- |
| Log<sub>10</sub>PRR<sub>48</sub> (95% CI) | 1.23 (1.12 - 1.33) | 1.51 (1.41 - 1.61) | 2.68 (2.46 - 2.89) | 3.42 (3.20 - 3.64) |
| PCt<sub>1/2</sub> [h (95% CI)] | 11.77 (10.84 - 12.87) | 9.57 (8.96 - 10.27) | 5.40 (5.00 - 5.87) | 4.23 (3.97 - 4.51) |
| Parasite regrowth incidence [n (%)] | 3 (100) | 3 (75) | 0 (0) | 0 (0) |

Log<sub>10</sub>PRR<sub>48</sub>: logarithm to base 10 of the parasite reduction ratio over a 48-hour period; PCt<sub>1/2</sub>: parasite clearance half-life. <sup>a</sup>One participant dosed with 200 mg tafenoquine had a non-significant regression fit and was therefore omitted from the PRR<sub>48</sub> and PCt<sub>1/2</sub> estimates.

**Figure 2.**
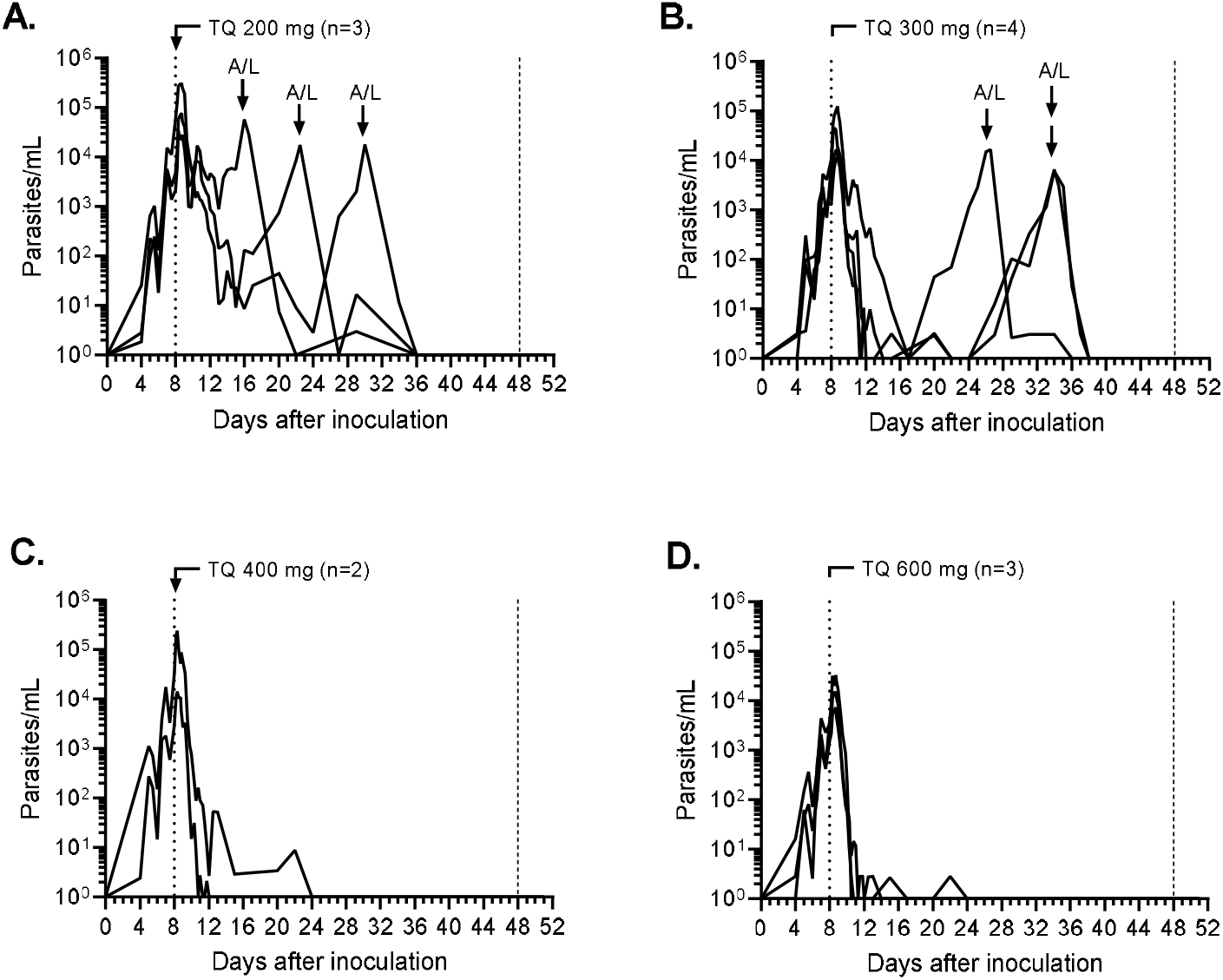
Individual participant parasitaemia-time profiles. Participants were inoculated intravenously with *P. falciparum*-infected erythrocytes on day 0 and were administered a single oral dose of 200 mg (A), 300 mg (B), 400 mg (C) or 600 mg (D) tafenoquine (TQ) on day 8 (indicated by the vertical dotted line). Definitive antimalarial treatment with a standard course of artemether-lumefantrine (A/L) was initiated in response to parasite regrowth or on day 48±2 (indicated by the vertical dashed line). Parasitaemia was measured using qPCR targeting the gene encoding *P. falciparum*18S rRNA.

### Tafenoquine exposure

Dose-related increases in tafenoquine exposure (maximum concentration [C_max_] and area under the concentration-time curve from time 0 to the last measured time point [AUC_0-last_]) were observed across the dose range (Table 3). Absorption was relatively slow, with time to maximum concentration (t_max_) approximately 12 h, while a long plasma elimination half-life (t_1/2_) of approximately 13 days was observed. Compared to plasma, venous whole blood tafenoquine exposure (C_max_ and AUC_0-last_) was higher across the dose range, while t_max_ and t_1/2_ were comparable (Table 3). Concentrations of the 5,6-orthoquinone metabolite were considerably lower than the parent compound in plasma (Figure S1), venous whole blood (Figure S2), and capillary whole blood (Figure S3), but substantially higher in urine (Figure S4).

**Table 3.** Tafenoquine plasma and venous whole blood non-compartmental pharmacokinetic parameters associated with single dose administration to healthy volunteers with induced *P. falciparum* parasitemia.

|  | Tafenoquine 200 mg<br>(N=3) |  | Tafenoquine 300 mg<br>(N=4) |  | Tafenoquine 400 mg<br>(N=2) |  | Tafenoquine 600 mg<br>(N=3) |  |
| --- | --- | --- | --- | --- | --- | --- | --- | --- |
|  | Plasma | Whole blood | Plasma | Whole blood | Plasma | Whole blood | Plasma | Whole blood |
| C<sub>max</sub> (ng/mL) | 138.2 (24.7) | 146.5 (40.9) | 205.8 (25.6) | 242.5 (30.2) | 324.8 (21.7) | 489.6 (24.1) | 340.2 (24.6) | 476.8 (35.3) |
| t<sub>max</sub> (h) | 12.2 (12.1 -<br>24.0) | 12.2 (12.1 -<br>48.1) | 12.0 (8.1 – 24.0) | 12.0 (8.0 -<br>12.0) | 10.0 (8.1 –<br>12.0) | 8.0 (8.0 - 8.1) | 12.1 (12.0 –<br>24.2) | 12.0 (8.0 -<br>24.2) |
| AUC<sub>0-last</sub> (h*ng/mL) | 47091 (9.2) | 53552 (17.0) | 67303 (33.4) | 83531 (28.4) | 117107 (25.0) | 158573 (22.5) | 128140 (28.2) | 157541 (33.7) |
| t<sub>1/2</sub> (h) | 309.4 (17.7) | 333.3 (5.0) | 357.3 (10.3) | 302.2 (14.2) | 313.9 (13.5) | 344.5 (14.6) | 314.4 (14.0) | 327.0 (4.7) |
| CL/F (L/h) | 3.8 (7.4) | 3.3 (13.6) | 3.9 (33.7) | 3.2 (26.9) | 3.1 (29.2) | 2.2 (26.5) | 4.1 (29.9) | 3.4 (33.4) |
| V<sub>z</sub>/F (L) | 1708 (22.0) | 1585 (18.2) | 2003 (33.5) | 1378 (34.0) | 1379 (15.2) | 1109 (11.6) | 1863 (24.6) | 1585 (30.8) |

Data are geometric means (coefficient of variation [%]) except t<sub>max</sub> which is median (range). C<sub>max</sub>: maximum observed concentration; t<sub>max</sub>: time to reach the maximum observed concentration; AUC<sub>0-last</sub>: area under the concentration-time curve from time 0 (dosing) to the last sampling time at which the concentration is at or above the lower limit of quantification; t<sub>1/2</sub>: apparent terminal half-life; CL/F: apparent total clearance; V<sub>z</sub>/F: apparent volume of distribution.

### Pharmacokinetic/Pharmacodynamic analysis

The PK of tafenoquine and 5,6-orthoquinone tafenoquine was described by a mammillary physiological model comprising seven disposition compartments (plasma, red blood cell and urine for both tafenoquine and 5,6-orthoquinone, and an additional tissue compartment for tafenoquine) with first-order absorption and linear elimination (Figure S5 and Table S1). Modelling identified time-dependent parasitemia as a significant covariate on tafenoquine clearance, resulting in decreasing clearance with increasing parasitemia. Visual predictive checks demonstrated that the observed profiles of tafenoquine fit well within the 5th and 95th percentiles of the simulated predictions, indicating the model adequacy in characterizing the observed data (Figure S6).

The relationship between tafenoquine red blood cell concentrations and parasite killing was described by an E_max_ PK/PD model. Modelling was unable to identify a significant relationship between 5,6-orthoquinone concentrations and parasite killing. Population PD parameters (Table S2) were estimated with satisfactory precision (relative standard error <30%). The inter-individual variability in the parasite growth rate constant was estimated with low precision and thus fixed to 0.07 based on retrospective *P. falciparum* parasite growth modelling in IBSM studies. Visual predictive checks demonstrated that the 5th, 50th and 95th percentiles of the observed data were within the 95% CI of the simulated data (Figure S7). PK/PD modelling using population estimates predicted an MIC of 74 ng/mL (0.16 nmol/mL) and a MPC_90_ of 561 ng/mL (1.21 nmol/mL) (Table S3 and Table S4).

### Tafenoquine dose predictions

The single doses of tafenoquine required to clear parasitaemia by a factor of 10^6^ and 10^9^, and to achieve APR_42_ with 90% probability, in all patients within a hypothetical endemic population was predicted using the PK/PD model with the associated inter-individual variability, covariate effect and model parameter uncertainty. Analyses were performed using body weights ranging from 5 to 60 kg, and baseline parasitemia log-normally distributed around a median of 10^7^ parasites/mL (inter-individual variability 60%), to simulate dosing an entire endemic population in the context of MDA. The predicted dose required to clear parasitemia by a factor of 10^6^ ranged from 50 mg (10 mg/kg) for a 5 kg infant to 460 mg (7.7 mg/kg) for a 60 kg adult (Table 4). Higher doses were predicted to be required to clear parasitemia by a factor of 10^9^ (eg. 65 mg for a 5 kg infant and 540 mg for a 60 kg adult), and to achieve APR_42_ (Table 4).

**Table 4.** Predicted single doses of tafenoquine required to clear asexual *P. falciparum* parasitemia in all patients within a hypothetical endemic population.

| Body weight (kg) | Dose to clear parasitemia by a factor of |  | Dose to achieve APR<sub>42</sub> with 90% probability<br>(mg) [mg/kg] |
| --- | --- | --- | --- |
|  | 10<sup>6</sup> (mg) [mg/kg] | 10<sup>9</sup> (mg) [mg/kg] |  |
| **5** | 50 [10] | 65 [13] | 90 [18] |
| **10** | 95 [9.5] | 120 [12] | 170 [17] |
| **15** | 140 [9.3] | 170 [11.3] | 260 [17.3] |
| **20** | 170 [8.5] | 210 [10.5] | 340 [17] |
| **30** | 250 [8.3] | 300 [10] | 480 [16] |
| **40** | 320 [8] | 380 [9.5] | 640 [16] |
| **50** | 390 [7.8] | 460 [9.2] | 780 [15.6] |
| **60** | 460 [7.7] | 540 [9] | 960 [16] |

Single doses of 1 to 40 mg/kg tafenoquine, with an increment of 1 mg/kg, were simulated in 20 trials of 100 patients with each body weight. Simulations were performed using the pharmacokinetic/pharmacodynamic model developed from the data obtained in healthy volunteers with induced blood stage *P. falciparum*. APR<sub>42</sub>: adequate parasitological response on day 42, defined as a decrease in parasitemia below 1 parasite in the body (or below the lower limit of quantification [LLOQ]) within 42 days, without early treatment failure (parasitemia above baseline at day 2 or above 25% of baseline at day 3) or late clinical failure (parasitemia above the LLOQ after Day 4).

### Safety and tolerability

A total of 160 AEs were reported, the majority (102/160) being mild to moderate signs and symptoms of malaria (Table 5). No AEs met the criteria of a serious adverse event or resulted in a participant discontinuing the study. There were 39 AEs considered related to tafenoquine (Table S5), with an increase in blood methemoglobin above the upper limit of normal (1.2%) the most commonly reported (10/12 participants). The peak methemoglobin concentration was related to the dose of tafenoquine administered, with 400 or 600 mg doses resulting in a greater elevation (2-5% at peak) compared to 200 or 300 mg doses (<2% at peak) (Figure S11). Blood methemoglobin AUC_0-last_ and C_max_ correlated with the AUC_0-last_ and C_max_ of tafenoquine and 5,6-orthoquinone in whole blood and plasma (Figures S12 and S13). The methemoglobin elevations were not accompanied by signs and symptoms associated with methemoglobinemia. A mild decrease in hemoglobin from baseline (pre-malaria inoculation) was recorded for 3 participants (largest decrease 24 g/L). Overall, hemoglobin concentrations did not appear to be dose related (Figure S10).

**Table 5.**
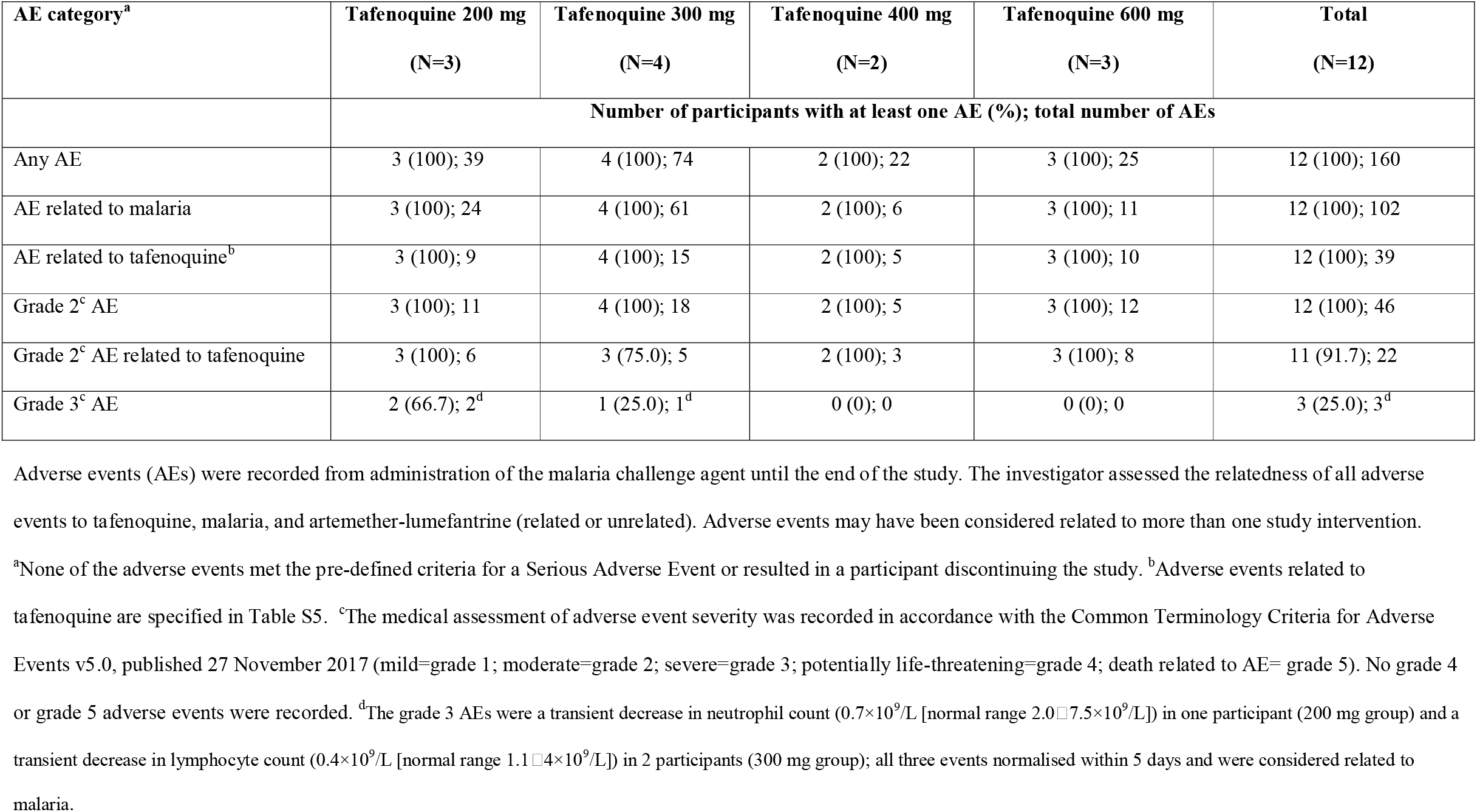
Summary of adverse events.

## DISCUSSION

The objective of this study was to characterize the blood stage *P. falciparum* antimalarial activity of tafenoquine in humans, with a view to potentially expanding the utility of the drug beyond its current indications of malaria prophylaxis and radical cure of vivax malaria. The fact that tafenoquine has a very long elimination half-life and exhibits activity against multiple parasite lifecycle stages, including the asexual blood stage [10, 11] and gametocytes responsible for transmission [21-23], indicated that it may be an appropriate candidate for MDA. The use of medicines with demonstrated efficacy for treatment, at full treatment doses, is important for MDA because some of the individuals treated will be parasitemic. It has been suggested that, at minimum, antimalarial drugs intended to clear asexual blood-stage parasitemia should clear parasites at least as fast as mefloquine, and reduce parasitemia by a factor of at least 10^6^ [24]. These properties served as benchmarks for our evaluation of tafenoquine.

The rate of parasite clearance with 400 mg or 600 mg tafenoquine (PCt_1/2_ 5.4 h and 4.2 h respectively) was faster than that previously characterized for 10 mg/kg mefloquine (PCt_1/2_ 6.2 h) using the IBSM model [25]. Doses of 200 mg or 300 mg tafenoquine were associated with a considerably slower rate of parasite clearance (PCt_1/2_ 11.8 h and 9.6 h respectively). Dose simulations in an endemic population took into account the higher parasite burdens, and greater variability in baseline parasitemia, compared to the volunteers in the current study. This was particularly important given that PK modelling revealed that higher parasitemia was associated with greater tafenoquine exposure due to reduced clearance. Adult doses of 460 mg (7.7 mg/kg) and 540 mg (9 mg/kg) were predicted to clear parasitemia by a factor of 10^6^ and 10^9^ respectively, while 960 mg (16 mg/kg) would achieve APR_42_ with 90% probability. The estimation of APR_42_ was considered informative albeit not entirely relevant for MDA, given that in practice all curative antimalarial treatments consist of combinations of two or more drugs, and/or require multiple doses.

The adult dose of tafenoquine predicted to clear asexual *P. falciparum* parasitaemia is higher than the dose currently recommended for preventing *P. vivax* relapse (300 mg), although it has recently been proposed that higher doses may be required to improve the effectiveness of anti-relapse treatment [26]. There is increased risk of *P. vivax* parasitemia following treatment of falciparum malaria in areas that are co-endemic for both species, and thus a rationale exists for administration of a radically curative treatment to patients with *P*.*falciparum*, even in the absence of detectable *P. vivax* [27]. Thus, the use of tafenoquine at the dose predicted to clear falciparum parasitaemia would likely also be beneficial in reducing the incidence of *P. vivax* relapse.

Although the results of this study indicate that tafenoquine exhibits potent blood stage antimalarial activity, the estimated doses required to clear asexual parasitemia will require prior screening for G6PD activity. A specific dose of tafenoquine that could be safely administered to G6PD-deficient individuals has not been established. However, a dose of 300 mg was found to result in a hemoglobin drop >2.5 g/dL in a small group of G6PD heterozygous adult females with enzyme activity 40-60% of normal [12]. Thus, it seems unlikely that the predicted doses required to clear asexual parasitemia could be safely administered for this purpose in the context of MDA. However, we and others have demonstrated that single low doses of tafenoquine (<100 mg adult dose) can reduce the transmission of *P. falciparum* to mosquitoes [22, 23]. Thus, the use of tafenoquine for this purpose, in combination with a partner drug to clear asexual parasitaemia, may be worthy of investigation for MDA.

As expected, no significant safety signals were associated with tafenoquine dosing of volunteers with normal G6PD levels in the current study. A mild transient decrease in hemoglobin occurred in a few participants, although this did not appear to be dose related and has been observed previously with blood stage malaria challenge of healthy volunteers [28]. A dose related transient increase in blood methemoglobin was observed, without clinical symptoms associated with methemoglobinemia. Similar observations have been reported following tafenoquine dosing of patients with *vivax* malaria [29-31]. The production of dose-dependent methemoglobinemia by 8-aminoquinoline antimalarials is associated with their efficacy against latent *P. vivax* and *P. ovale* [32]. The results of our study suggest that a similar phenomenon may exist for tafenoquine activity against asexual blood stage *P. falciparum*. Doses of 400 or 600 mg resulted in a notably greater elevation in methemoglobin compared to doses of 200 or 300 mg, while also resulting in faster parasite clearance without subsequent parasite regrowth. The specific mechanisms of tafenoquine-induced parasite killing, hemolytic toxicity, and methemoglobinemia remain unclear. It is hypothesized that tafenoquine exhibits parasite stage-specific activity associated with different metabolites and different modes of action [33]. Activity against blood stage parasites is thought to be independent of CYP-2D6 metabolism, potentially involving interference with heme polymerization, or oxidative mechanisms [33]. The 5,6-orthoquinone metabolite of primaquine is considered a surrogate marker for active metabolites of primaquine [34]. Although we were able to measure the 5,6-orthoquinone metabolite of tafenoquine, a relationship between the concentration of this metabolite and parasite killing was not observed. Further work to better understand the role of the metabolite and the mode of action of tafenoquine is warranted.

The primary limitation of this study is that the predicted doses required to clear parasitemia in an endemic population were based on the assumption that the PK/PD relationship characterized in the volunteers would be the same in the target population. We cannot discount the possibility that the PK/PD relationship between tafenoquine and parasite killing would be influenced by other factors such as host immunity, G6PD activity, and other bioactive metabolites. However, estimates of therapeutic doses of other antimalarial drugs in volunteers has translated well to patient populations [25, 35].

In conclusion, this study has demonstrated that tafenoquine exhibits potent *P. falciparum* blood stage antimalarial activity in humans when administered as a single dose. However, the predicted doses required to clear asexual parasitemia will require prior screening for G6PD activity. This would limit its use for the purpose of clearing asexual parasitaemia in MDA where widespread G6PD screening would prove costly and logistically difficult.

## Data Availability

All data produced in the present study are available upon reasonable request to the authors.

## Abbreviations

AEs: adverse events
G6PD: glucose 6-phosphate dehydrogenase
IBSM: induced blood stage malaria
MDA: mass drug administration
PK/PD: pharmacokinetic/pharmacodynamics

## ACKNOWLEWDGEMENTS

We thank all the volunteers who participated in the study, staff at the University of the Sunshine Coast Clinical Trials Unit who conducted the trial, staff at the Queensland Paediatric Infectious Diseases laboratory for qPCR analysis, Dr. Scott Miller from the Bill and Melinda Gates Foundation for valuable discussions as part of the Safety and Data Review Team, and Karin Van Breda from the Australian Defence Force Malaria and Infectious Disease Institute for the measurement of tafenoquine concentrations in biospecimens.

## DISCLAIMER

The views expressed in this article are those of the authors and do not necessarily reflect the official policy or position of the Australian Defence Force, Joint Health Command or any extant Australian Defence Force policy.

## SUPPLEMENTARY MATERIAL

- Figure S1. Individual participant plasma concentrations of tafenoquine and 5, 6-orthoquinone over time per dose.
- Figure S2. Individual participant venous whole blood concentrations of tafenoquine and 5, 6-orthoquinone over time per dose.
- Figure S3. Individual participant capillary whole blood concentrations of tafenoquine and 5, 6-orthoquinone over time per dose.
- Figure S4. Cumulative amount of tafenoquine and 5, 6-orthoquinone excreted in the urine for each participant over time per dose.
- Text S1. Pharmacokinetic/pharmacodynamic analysis extended methods.
- Figure S5. Schematic diagram of the population pharmacokinetic/pharmacodynamic model of tafenoquine and 5, 6-orthoquinone.
- Table S1. Population parameter estimates of the final tafenoquine pharmacokinetic model.
- Figure S6. Visual predictive checks of the population pharmacokinetic model of tafenoquine and 5, 6-orthoquinone.
- Table S2. Population estimates of the final pharmacokinetic/pharmacodynamic model.
- Figure S7. Visual predictive checks of the population pharmacokinetic/pharmacodynamic model of tafenoquine and 5, 6-orthoquinone.
- Table S3. Efficacy parameters based on population and individual estimates following administration of single doses of tafenoquine succinate.
- Table S4. Corresponding tafenoquine population pharmacodynamic parameters in different biological matrices.
- Figure S8. Simulated total parasite reduction ratio in a hypothetical patient population following administration of different single doses of tafenoquine.
- Figure S9. Probability of adequate parasitological response within 42 days (APR_42_) for different single doses of tafenoquine.
- Table S5. Incidence of adverse events related to tafenoquine.
- Figure S10. Individual participant hemoglobin-time profiles.
- Figure S11. Individual participant methemoglobin-time profiles.
- Figure S12. Correlation between blood methemoglobin exposure and tafenoquine and 5,6-orthoquinone exposure in whole blood and plasma.
- Figure S13. Correlation between maximum blood methemoglobin concentration and maximum tafenoquine and 5,6-orthoquinone concentrations in whole blood and plasma.
- Text S2. Participant eligibility criteria.

