## Supplementary material for "Characterizing the blood stage antimalarial activity of tafenoquine in healthy volunteers experimentally infected with *Plasmodium falciparum*"

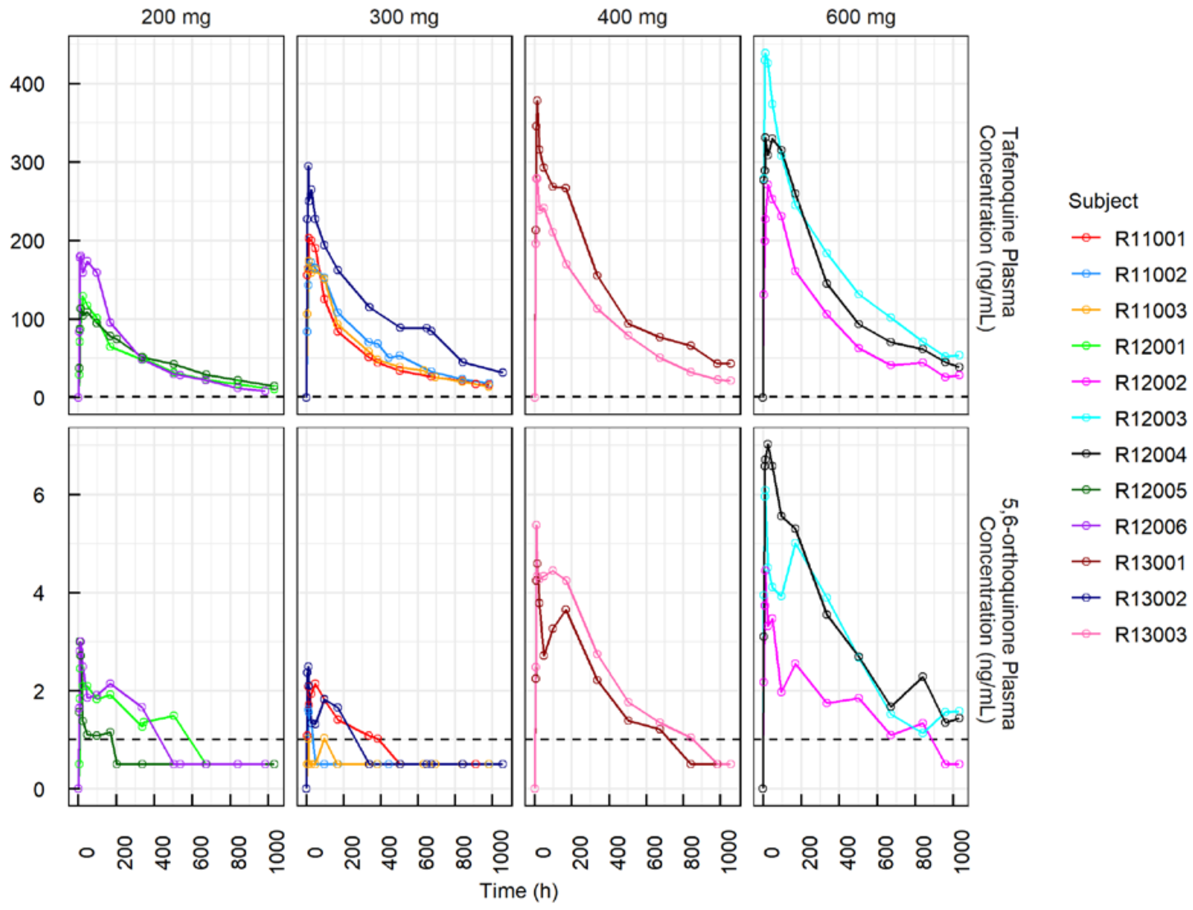

**Figure S1. Individual participant plasma concentrations of tafenoquine and 5, 6-orthoquinone over time per dose**

Dashed lines represent the lower limit of quantification.

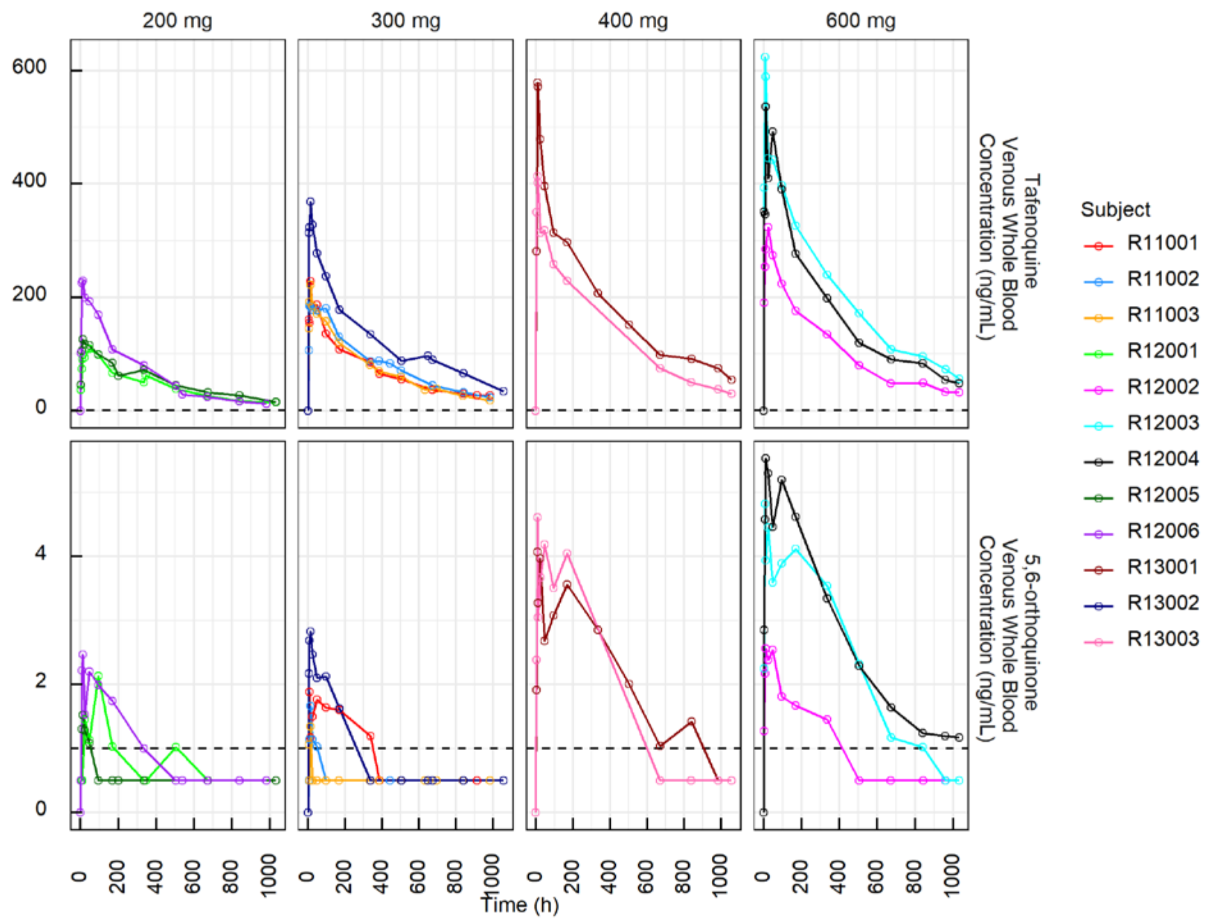

**Figure S2. Individual participant venous whole blood concentrations of tafenoquine and 5, 6-orthoquinone over time per dose**

Dashed lines represent the lower limit of quantification.

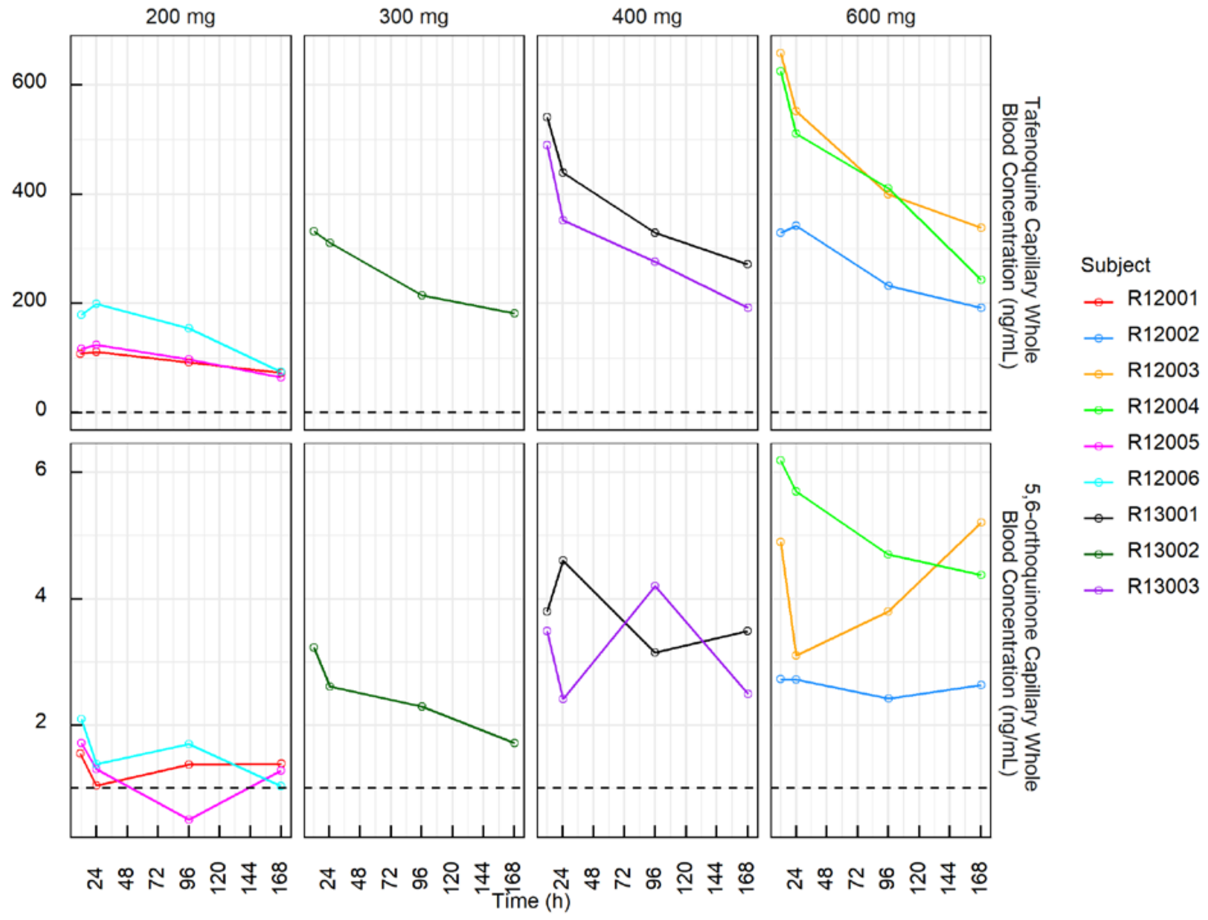

**Figure S3. Individual participant capillary whole blood concentrations of tafenoquine and 5, 6-orthoquinone over time per dose**

Dashed lines represent the lower limit of quantification. Capillary blood was not collected for cohort 1 (n=3 participants dosed with 300 mg).

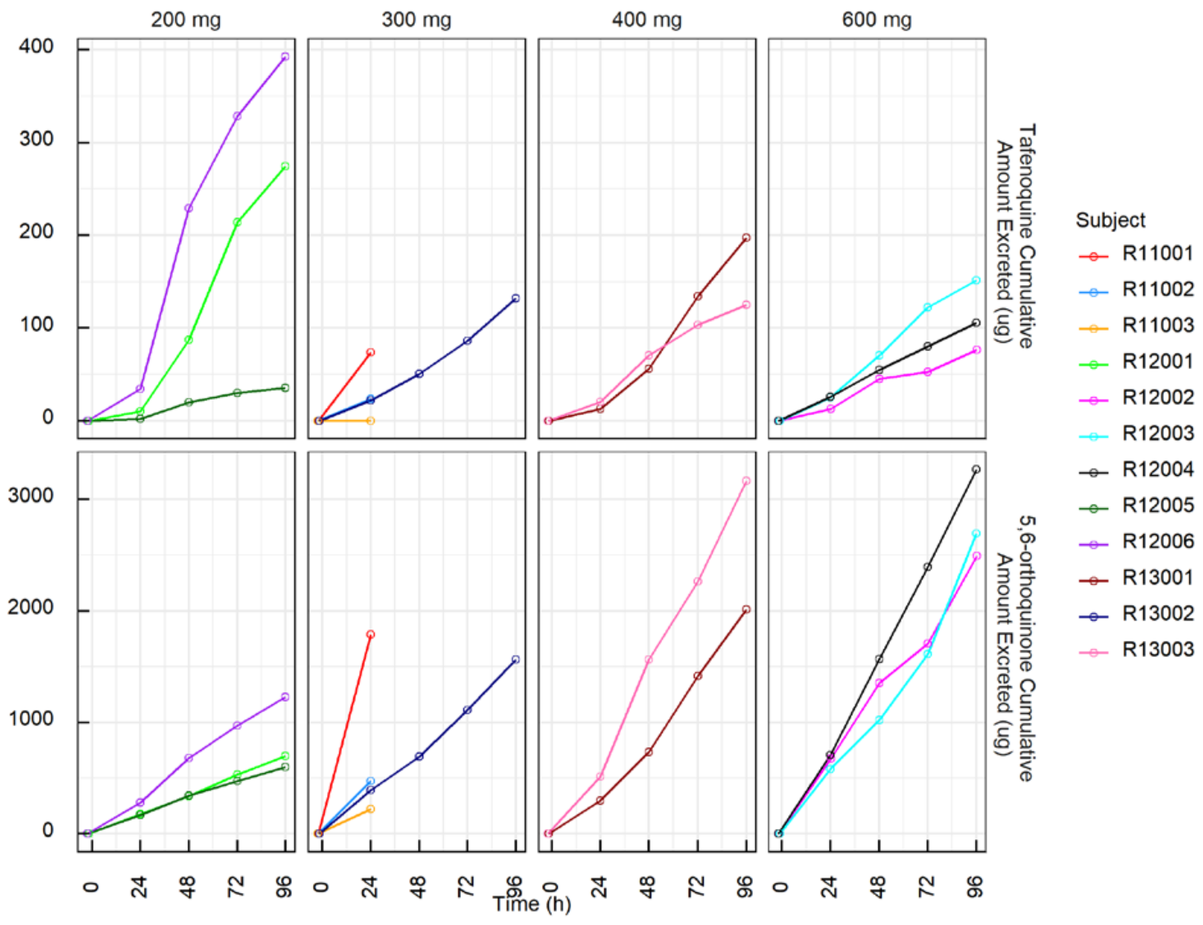

**Figure S4. Cumulative amount of tafenoquine and 5, 6-orthoquinone excreted in the urine for each participant over time per dose**

Urine was only collected up to 24 hours post-dose for cohort 1 (n=3 participants dosed with 300 mg).

**Text S1. Pharmacokinetic/pharmacodynamic analysis extended methods**

Model selection

Throughout the population PK/PD model building, hierarchical models were compared based on the likelihood ratio test (difference in objective function value, ΔOFV) in which improvement of model fit between models was determined using chi-squared distribution with one degree of freedom (df). Non-hierarchical models were compared using the Bayesian information criterion (BIC).

Population PK modelling

The tafenoquine succinate doses were converted into molar unit using the molecular mass of 581.6 g/mol. Tafenoquine and 5, 6-orthoquinone plasma and venous and capillary whole blood concentrations were converted into molar unit using the molecular masses of 463.5 g/mol and 303.4 g/mol, respectively. Tafenoquine and 5, 6-orthoquinone cumulative amount excreted in the urine were calculated by multiplying their urine concentrations with the urine volume and converted into molar unit. Data below the lower limit of quantification (LLOQ) were treated using left-censoring method, by maximising the likelihood of these data reported of being below LLOQ.

Population PK modelling was performed in several steps. Initially, an exploratory analysis with the tafenoquine and 5, 6-orthoquinone plasma concentration-time profiles (alone and combined) was undertaken to investigate a base structural model. One-, two- and three-compartment with different absorption and elimination models were tested. Zero- and first-order, and transit compartment absorption models with or without lag time were examined. Linear, saturable and parallel linear and saturable elimination were also evaluated. During this step, a complete and irreversible *in vivo* conversion of tafenoquine into 5, 6-orthoquinone was assumed to avoid identifiability problem. Catenary and mammillary structural arrangements were then explored to simultaneously model the plasma and venous whole blood concentration-time profiles of tafenoquine and 5, 6-orthoquinone. Proportional conversion factors were estimated to fit the venous and capillary whole blood concentrations of tafenqouine and 5, 6-orthoquinone simultaneously. The PK model was subsequently expanded by incorporating cumulative amount of tafenoquine and 5, 6-orthoquinone in the urine data. Interindividual variability (IIV) of the PK parameters was assumed to be normally distributed on log-scale.

Covariate analysis was performed to explore the source of variability in the models using the stepwise covariate modelling approach. The covariate effects were considered based on biological plausibility, statistical significance and clinical relevance. The correlation between covariates was explored to identify any highly correlated covariates. The parameter-covariate relationships were identified based on empirical Bayes estimates (parameter's ETA) versus covariates plots and were considered in covariate model building if the parameter's ETA-covariate correlation was larger than 0.3 for continuous covariates, or the parameter's ETA was significantly different as tested by t-test between categorical covariates. The relationship of model parameter and potential covariates (*i.e.* age, gender, tafenoquine succinate dose, baseline parasitemia, time-dependent parasitemia and cytochrome P450 (CYP) 2D6 genetic polymorphism) were evaluated by stepwise forward inclusion ($p < 0.05$, ΔOFV = -3.84) and backward exclusion ($p < 0.001$, ΔOFV = -10.83) using linear, power, exponential and $E_{max}$ functions. Body weight was evaluated as an allometric function on apparent clearance (scaled to the power of 0.75) and volume of distribution parameters (scaled to the power of 1) and was centred on 60 kg.

Population PK/PD modelling

Each *P. falciparum* parasitemia sample was quantified in triplicate per time-point and natural log (ln)-transformed. The mean of the ln-transformed parasitemia was used as a summary parasitemia value per time-point per participant.

A sequential PK/PD modelling approach was used in which the empirical Bayes estimates of individual PK parameters were used as regression parameters for the PK/PD model. The observations below the limit of detection (LOD) were modelled using a left-censoring method. In this analysis, the changes in parasitemia over time in the presence of antimalarial drug are assumed to be a result of the difference between parasite growth rate ($k_{grow}$) and the rate at which parasites are killed by the drug ($k_{kill}$) with a baseline parasitemia ($PL_{base}$) at time $t_0$ (time of first observation).

Different structural PK/PD models were tested to assess the relationship of tafenoquine and/or 5, 6-orthoquinone red blood cell concentration on the parasite killing rate. The $E_{max}$ model assumed a direct effect of tafenoquine and/or 5, 6-orthoquinone red blood cell concentrations on parasite killing rate. The turnover model assumed an indirect effect of tafenoquine and/or 5, 6-orthoquinone red blood cell concentrations on parasite killing rate due to lag in the PD processes (*i.e.* the kinetics of drug-receptor binding). The effect compartment model assumed an indirect effect of tafenoquine and/or 5, 6-orthoquinone red blood cell concentrations on parasite killing rate due to biophase equilibration. This model can be interpreted as the delay for the analyte to reach site of action in which the tafenoquine and/or 5, 6-orthoquinone concentrations in the effect compartment drives the parasite killing effect. The parasite clearance model assumed that the observed parasites were a mixture of living and dead parasites and that the tafenoquine and/or 5, 6-orthoquinone red blood cell concentrations had a direct effect on the killing rate of the parasites. The competitive binding model was used to describe the relationship of tafenoquine and 5, 6-orthoquinone on parasite killing. It was assumed that both tafenoquine and 5, 6-orthoquinone are proficient of binding to the same binding site.

PD parameters were modelled assuming a log-normal distribution with exception of *ln*-transformed baseline parasite values where a normal distribution was assumed. An exponential random effect was used for IIV in PD parameters. Similarly, IIV was initially included in all PD parameters; it was fixed to zero if its inclusion was not supported by the data. RUV was modelled as an additive error on the logarithmically transformed observations which corresponded to an exponential error on an arithmetic scale. Four estimations were made for the PD parameters with randomised initial values.

Calculation of key efficacy parameters

The minimum inhibitory concentration (MIC), the minimal parasiticidal concentration ($MPC_{90}$) and the parasite reduction rate within 48 hours post-dose ($PRR_{48}$) were derived from the PD models. MIC was defined as the lowest drug concentration which inhibited parasite growth. $MPC_{90}$ was defined as the lowest analyte concentration needed at which the parasite killing rate was equal to 90% of its maximum.

Definition of adequate parasitological response on day 42 ($APR_{42}$)

$APR_{42}$ was defined as a decrease in parasitemia below the LLOQ or below 1 parasite in the body within 42 days and did not meet any criteria for early treatment failure (ETF) or late clinical failure (LCF). ETF was defined as parasitemia above baseline at Day 2 or above 25% of baseline at Day 3. LCF was defined as parasitemia above the LLOQ after Day 4.

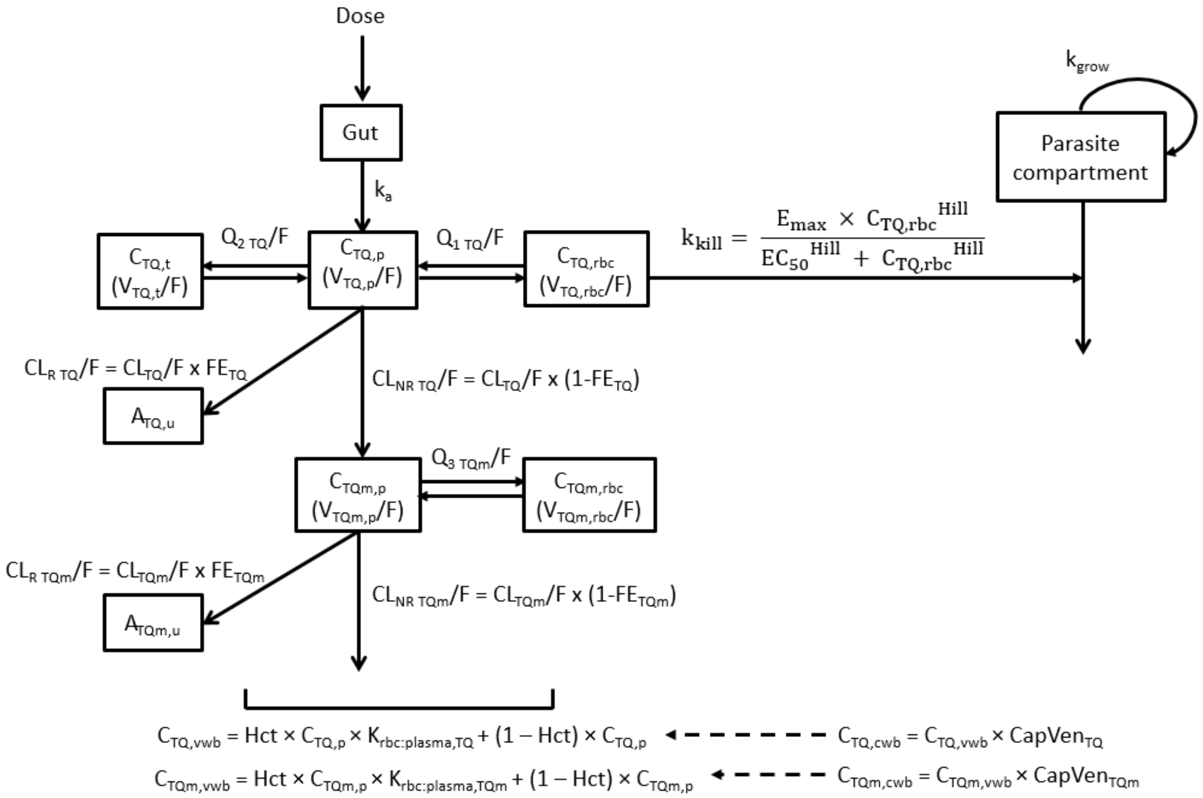

**Figure S5. Schematic diagram of the population pharmacokinetic/pharmacodynamic model of tafenoquine and 5, 6-orthoquinone**

TQ = tafenoquine, TQm = 5, 6-orthoquinone, $C_{(analyte),p}$ = analyte plasma concentration, $C_{(analyte),rbc}$ = analyte red blood cell concentration, $C_{(analyte),t}$ = analyte tissue concentration, $C_{(analyte),vwb}$ = analyte venous whole blood concentration, $C_{(analyte),cwb}$ = analyte capillary whole blood concentration, $A_{(analyte),u}$ = analyte cumulative urinary amount, $k_a$ = rate constant of first-order absorption, F = relative bioavailability of first-order absorption, $CL_{(analyte),}$/F = apparent clearance of analyte, $CL_{R\ (analyte)}$/F = apparent renal clearance of analyte, $CL_{NR\ (analyte)}$/F = apparent non-renal clearance of analyte, $V_{(analyte),\ p}$/F = apparent volume of distribution of analyte in plasma compartment, $V_{(analyte),\ rbc}$/F = apparent volume of distribution of analyte in red blood cell compartment, $V_{(analyte),\ t}$/F = volume of distribution of analyte in tissue compartment, $Q_{1\ TQ}$/F = apparent inter-compartmental tafenoquine clearance between plasma and tissue compartments, $Q_{2\ TQ}$/F = apparent inter-compartmental tafenoquine clearance between plasma and red blood cell compartments, $Q_{3\ TQm}$/F = apparent inter-compartmental 5, 6-orthoquinone clearance between plasma and red blood cell compartments, $CL_{R\ TQ}$/F = apparent tafenoquine renal clearance, $CL_{R\ TQm}$/F = apparent 5, 6-orthoquinone renal clearance, $CL_{NR\ TQ}$/F = apparent tafenoquine non-renal clearance, $CL_{NR\ TQm}$/F = apparent 5, 6-orthoquinone non-renal clearance, $FE_{TQ}$ = fraction of tafenoquine urinary excretion, $FE_{TQm}$ = fraction of 5, 6-orthoquinone urinary excretion, $CapVen_{(analyte)}$ = capillary-venous conversion factor of analyte, Hct = time-varying haematocrit, $K_{rbc:plasma,\ (analyte)}$ = red blood cell-to-plasma partition coefficient, $k_{grow}$ = growth rate of *P. falciparum*, $k_{kill}$ = killing rate of *P. falciparum* by the drug, $E_{max}$ = maximum parasite killing attributable to the analyte, $EC_{50}$ = the analyte concentration producing 50% of the $E_{max}$, Hill = the Hill coefficient, $C_{TQ,\ rbc}$ = tafenoquine red blood cell concentration.

**Table S1. Population parameter estimates of the final tafenoquine pharmacokinetic model**

| Parameter | Value | RSE | Shrinkage | Comment |
|---|---|---|---|---|
| F | 1 (fixed) | - | - | Relative bioavailability (-) |
| $k_a$ | 0.222 | 12.3 | - | Absorption rate constant (/h) |
| $CL_{TQ}/F$ | 2.8 | 5.8 | - | Apparent tafenoquine clearance (L/h) |
| $V_{TQ, p}/F$ | 876 | 5.3 | - | Apparent volume of distribution of tafenoquine in plasma compartment (L) |
| $Q_{1\ TQ}/F$ | 4.15 | 1.6 | - | Apparent inter-compartmental tafenoquine clearance between plasma and tissue compartments (L/h) |
| $V_{TQ, t}/F$ | 341 | 9.1 | - | Apparent volume of distribution of tafenoquine in tissue compartment (L) |
| $Q_{2\ TQ}/F$ | 1.07 | 0.01 | - | Apparent inter-compartmental tafenoquine clearance between plasma and red blood cell compartments (L/h) |
| $V_{TQ, rbc}/F$ | 0.0469 | 3.1 | - | Apparent volume of distribution of tafenoquine in red blood cell compartment (L) |
| $K_{rbc:plasma, TQ}$ | 1.6 | 4.6 | - | Tafenoquine red blood cell-to-plasma partition coefficient |
| $CapVen_{TQ}$ | 1.01 | 2.5 | - | Tafenoquine capillary-venous whole blood conversion factor |
| $FE_{TQ}$ | 0.00562 | 17.0 | - | Fraction of tafenoquine excreted in the urine |
| $CL_{TQm}/F$ | 97.4 | 10.9 | - | Apparent 5, 6-orthoquinone clearance (L/h) |
| $V_{TQm, p}/F$ | 51.2 | 4.7 | - | Apparent volume of distribution of 5, 6-orthoquinone in plasma compartment (L) |
| $Q_{3\ TQm}/F$ | 0.381 (fixed) | - | - | Apparent inter-compartmental 5, 6-orthoquinone clearance between plasma and red blood cell compartments (L/h) |
| $V_{TQm, rbc}/F$ | 30.1 (fixed) | 3.1 | - | Apparent volume of distribution of 5, 6-orthoquinone in red blood cell compartment (L) |
| $K_{rbc:plasma, TQm}$ | 0.472 | 27.2 | - | 5, 6-orthoquinone red blood cell-to-plasma partition coefficient |
| $CapVen_{TQm}$ | 1.02 | 4.4 | - | 5, 6-orthoquinone capillary-venous whole blood conversion factor |
| $FE_{TQm}$ | 0.0427 | 13.6 | - | Fraction of 5, 6-orthoquinone excreted in the urine |
| **Inter-individual variability[a]** | | | | |
| $\omega_{k_a}$ | 0.392 | 23.5 | 3.8 | Log-normal |
| $\omega_{CL_{TQ}/F}$ | 0.196 | 21.3 | -2.4 | Log-normal |
| $\omega_{V_{TQ,p}/F}$ | 0.172 | 23.4 | 2.4 | Log-normal |
| $\omega_{V_{TQ,t}/F}$ | 0.231 | 37.0 | 21.2 | Log-normal |
| $\omega_{K_{rbc:plasma,TQ}}$ | 0.126 | 31.0 | 14.6 | Log-normal |
| $\omega_{FE_{TQ}}$ | 0.517 | 25.7 | 9.6 | Log-normal |
| $\omega_{CL_{TQm}/F}$ | 0.368 | 21.7 | -1.6 | Log-normal |
| $\omega_{K_{rbc:plasma,TQm}}$ | 0.738 | 30.7 | 17.6 | Log-normal |
| $\omega_{FE_{TQm}}$ | 0.482 | 22.0 | -2.6 | Log-normal |

| | | | | |
|---|---|---|---|---|
| **Parameter-covariate relationship** | | | | |
| $\beta_{CL_{TQ}/F,WT}$[b] | 0.75 (fixed) | - | - | Body weight on $CL_{TQ}/F$ |
| $\beta_{V_{TQ,p}/F,WT}$[b] | 1 (fixed) | - | - | Body weight on $V_{TQ,\ p}/F$ |
| $\beta_{Q_{1\ TQ}/F,WT}$[b] | 0.75 (fixed) | - | - | Body weight on $Q_{1\ TQ}/F$ |
| $\beta_{V_{TQ,t}/F,WT}$[b] | 1 (fixed) | - | - | Body weight on $V_{TQ,\ t}/F$ |
| $\beta_{Q_{2\ TQ}/F,WT}$[b] | 0.75 (fixed) | - | - | Body weight on $Q_{2\ TQ}/F$ |
| $\beta_{V_{TQ,rbc}/F,WT}$[b] | 1 (fixed) | - | - | Body weight on $V_{TQ,\ rbc}/F$ |
| $\beta_{CL_{TQm}/F,WT}$[b] | 0.75 (fixed) | - | - | Body weight on $CL_{TQm}/F$ |
| $\beta_{V_{TQm,p}/F,WT}$[b] | 1 (fixed) | - | - | Body weight on $V_{TQm,\ p}/F$ |
| $\beta_{Q_{3\ TQm}/F,WT}$[b] | 0.75 (fixed) | - | - | Body weight on $Q_{3\ TQm}/F$ |
| $\beta_{V_{TQm,rbc}/F,WT}$[b] | 1 (fixed) | - | - | Body weight on $V_{TQm,\ rbc}/F$ |
| $\beta_{CL_{TQ}/F,PARA}$[c] | -0.0355 (fixed) | - | - | Time-dependent parasitemia on $CL_{TQ}/F$ |
| $\beta_{FE_{TQ},DOSE}$[d] | -1.12 (fixed) | - | - | Dose on $FE_{TQ}$ |
| $DOSE_{max,FE_{TQm}}$[e] | 0.266 (fixed) | - | - | Maximum effect of dose on $FE_{TQm}$ |
| $DOSE_{50,FE_{TQm}}$[e] | 0.205 (fixed) | - | - | Dose which produces 50% of the maximum covariate effect of $FE_{TQm}$ |
| **Residual unexplained variability** | | | | |
| $\varepsilon_{prop,TQ,p}$ | 0.126 | 6.9 | 6.2 | Proportional error model for tafenoquine plasma data |
| $\varepsilon_{prop,TQm,p}$ | 0.254 | 8.4 | 4.8 | Proportional error model for 5, 6-orthoquinone plasma data |
| $\varepsilon_{prop,TQ,vwb}$ | 0.119 | 6.6 | 6.3 | Proportional error model for tafenoquine venous whole blood data |
| $\varepsilon_{prop,TQm,vwb}$ | 0.225 | 9.2 | 9.2 | Proportional error model for 5, 6-orthoquinone venous whole blood data |
| $\varepsilon_{prop,TQ,cwb}$ | 0.125 | 13.3 | 3.2 | Proportional error model for tafenoquine capillary whole blood data |
| $\varepsilon_{prop,TQm,cwb}$ | 0.212 | 13.5 | 2.9 | Proportional error model for 5, 6-orthoquinone capillary whole blood data |
| $\varepsilon_{add,TQ,u}$ | 0.00043 | 13.3 | 14.6 | Additive error model for tafenoquine urine data |
| $\varepsilon_{prop,TQm,u}$ | 0.254 | 8.4 | 4.8 | Proportional error model for 5, 6-orthoquinone urine data |

RSE = relative standard error.

[a] Standard deviation of inter-individual variability.

[b] ΔOFV = -14.79 (0 df).

[c] $CL_{TQ}/F \times \exp^{\left(\beta_{CL_{TQ}/F,PARA} \times \left(\frac{PARA_i}{0.9632}\right)\right)}$ where $PARA_i$ was individual time-dependent parasitemia (ΔOFV = -44.37; 1 df).

[d] $FE_{TQ} \times \left(1 + \beta_{FE_{TQ},DOSE} \times (DOSE_i - 0.3439)\right)$ where $DOSE_i$ was individual dose (ΔOFV = -14.16; 1 df).

[e] $FE_{TQm} \times \left(1 + \left(\frac{(DOSE_i - 0.3439) \times DOSE_{max,FE_{TQm}}}{(DOSE_i - 0.3439) + \left(DOSE_{50,FE_{TQm}} - 0.3439\right)}\right)\right)$ where $DOSE_i$ was individual dose (ΔOFV = -13.13; 2df).

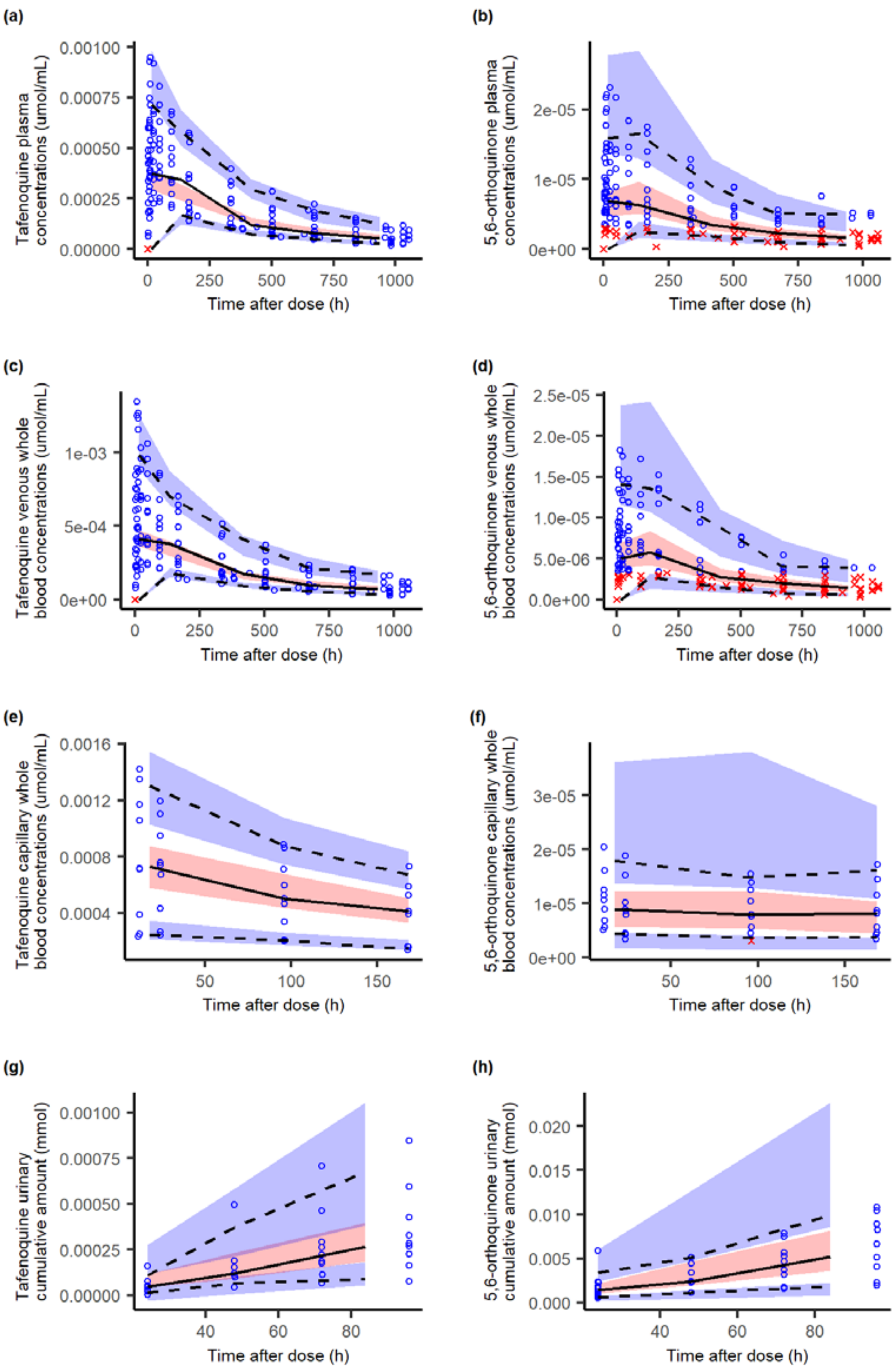

**Figure S6. Visual predictive checks of the population pharmacokinetic model of tafenoquine and 5, 6-orthoquinone**

Blue circles represent observed data; red crosses represent simulated data below the lower limit of quantification; lines represent the 5th, 50th and 95th percentiles for the observed data; blue shaded areas represent the confidence intervals for 5th and 95th percentiles; pink shaded areas represent the 95% confidence interval for the 50th percentile; derived from 1000 stochastic profiles simulated from the final population pharmacokinetic model.

**Table S2. Population estimates of the final pharmacokinetic/pharmacodynamic model**

| Parameter | Value | RSE | Shrinkage | Comment |
|---|---|---|---|---|
| $PL_{base}$ | -7.04 | 14.0 | - | Log-transformed baseline parasitemia (parasites/mL) |
| $k_{grow}$ | 0.0904 | 6.6 | - | Parasite growth rate (/h) |
| $E_{max}$ | 0.282 | 6.4 | - | Maximum parasite clearance rate (/h) |
| $EC_{50}$ | 0.268 | 18.2 | - | Concentration achieving 50% of maximum effect (nmol/mL) |
| Hill | 1.46 | 11.7 | - | Hill coefficient (-) |
| **Inter-individual variability[a]** | | | | |
| $\omega_{k_{grow}}$ | 0.07 (fixed) | - | 19.9 | Log-normal |
| $\omega_{EC_{50}}$ | 0.202 | 27.8 | 8.9 | Log-normal |
| $\omega_{Hill}$ | 0.2 (fixed) | - | 0.5 | Log-normal |
| **Residual variability** | | | | |
| $\varepsilon_{add}$ | 1.21 | 4.3 | 7.0 | Log-transformed parasitemia (parasites/mL) |

RSE = relative standard error.

[a]Standard deviation of inter-individual variability.

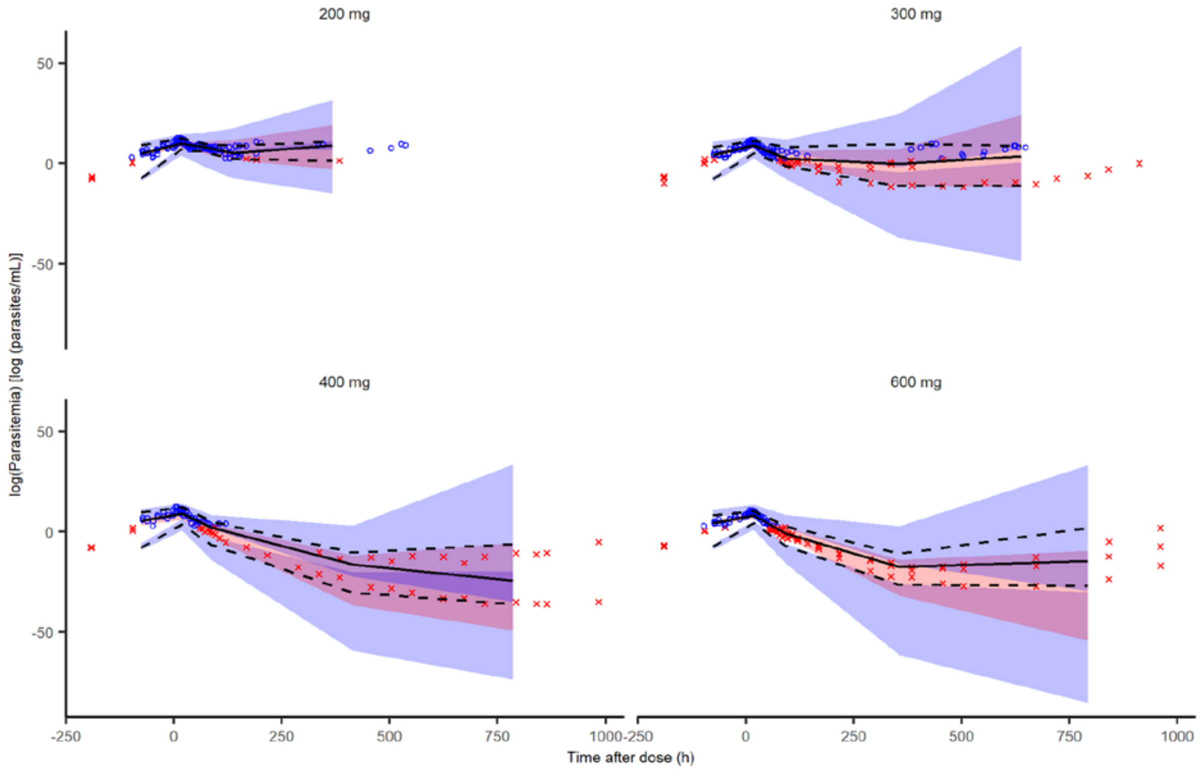

**Figure S7. Visual predictive checks of the population pharmacokinetic/pharmacodynamic model of tafenoquine and 5, 6-orthoquinone**

Blue circles represent the observed data; red crosses represent data simulated below the lower limit of quantification; lines represent the 5th, 50th and 95th percentiles for the observed data; blue shaded areas represent the confidence intervals for 5th and 95th percentiles; pink shaded areas represent the 95% confidence interval for the 50th percentile; derived from 1000 stochastic profiles simulated from the final population pharmacokinetic/pharmacodynamic model. Log refers to natural-logarithm.

**Table S3. Efficacy parameters based on population and individual estimates following administration of single doses of tafenoquine succinate**

| Parameter | Population estimate | Individual estimate[a] |
|---|---|---|
| MIC (nmol/mL) | 0.16 | 0.10 – 0.27 |
| MIC (ng/mL) | 74 | 46 – 123 |
| $MPC_{90}$ (nmol/mL) | 1.21 | 0.70 – 1.84 |
| $MPC_{90}$ (ng/mL) | 562 | 322 – 854 |
| $PRR_{48}$ ($log_{10}$ unit) | 4.00 | 3.78 – 4.11 |
| Parasite clearance half-life (h) | 3.61 | 3.51 – 3.82 |
| Time above MIC (days) | | |
| 200 mg | 13 | 3 – 13 |
| 300 mg | 24 | 10 – 28 |
| 400 mg | 32 | 29 – 43 |
| 600 mg | 43 | 30 – 44 |

MIC = minimum inhibitory concentration (*i.e.* tafenoquine red blood concentration), $MPC_{90}$ = minimum parasiticidal concentration that at 90% of maximum effect (*i.e.* tafenoquine red blood cell concentration), $PRR_{48}$ = parasite reduction ratio within 48 hours. [a]Individual estimates are in minimum – maximum.

**Table S4. Corresponding tafenoquine population pharmacodynamic parameters in different biological matrices**

| Parameter | Plasma | Venous whole blood | Capillary whole blood |
|---|---|---|---|
| $EC_{50}$ (nmol/mL) | 0.268 | 0.43 | 0.432 |
| $EC_{50}$ (ng/mL) | 124 | 199 | 200 |
| MIC (nmol/mL) | 0.16 | 0.257 | 0.259 |
| MIC (ng/mL) | 74 | 119 | 120 |
| $MPC_{90}$ (nmol/mL) | 1.21 | 1.94 | 1.95 |
| $MPC_{90}$ (ng/mL) | 561 | 899 | 904 |

$EC_{50}$ = analyte concentration that produce half of the $E_{max}$, MIC = minimum inhibitory concentration, $MPC_{90}$ = minimum parasiticidal concentration that at 90% of maximum effect.

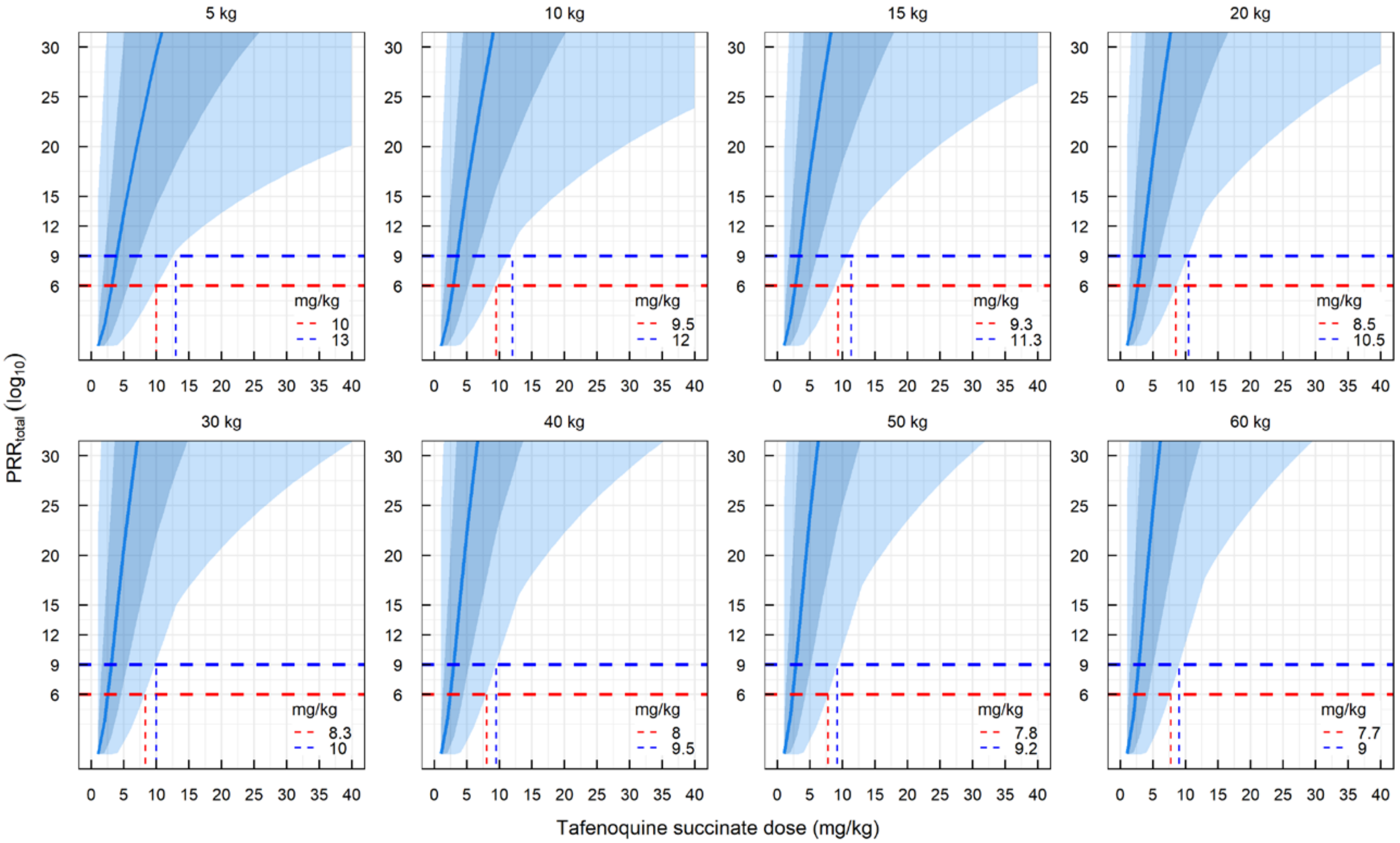

**Figure S8. Simulated total parasite reduction ratio in a hypothetical patient population following administration of different single doses of tafenoquine.** The blue solid line represents the median $PRR_{total}$; the dark blue shaded area represents the 5th-95th percentile; the light blue shaded area represents the 0-100th percentile (minimum-maximum); the red and blue dashed lines represent $PRR_{total}$ of 6- and 9-$log_{10}$, respectively. For each body weight, simulations were performed in 20 trials of 100 hypothetical patients each. $PRR_{total}$: total parasite reduction ratio.

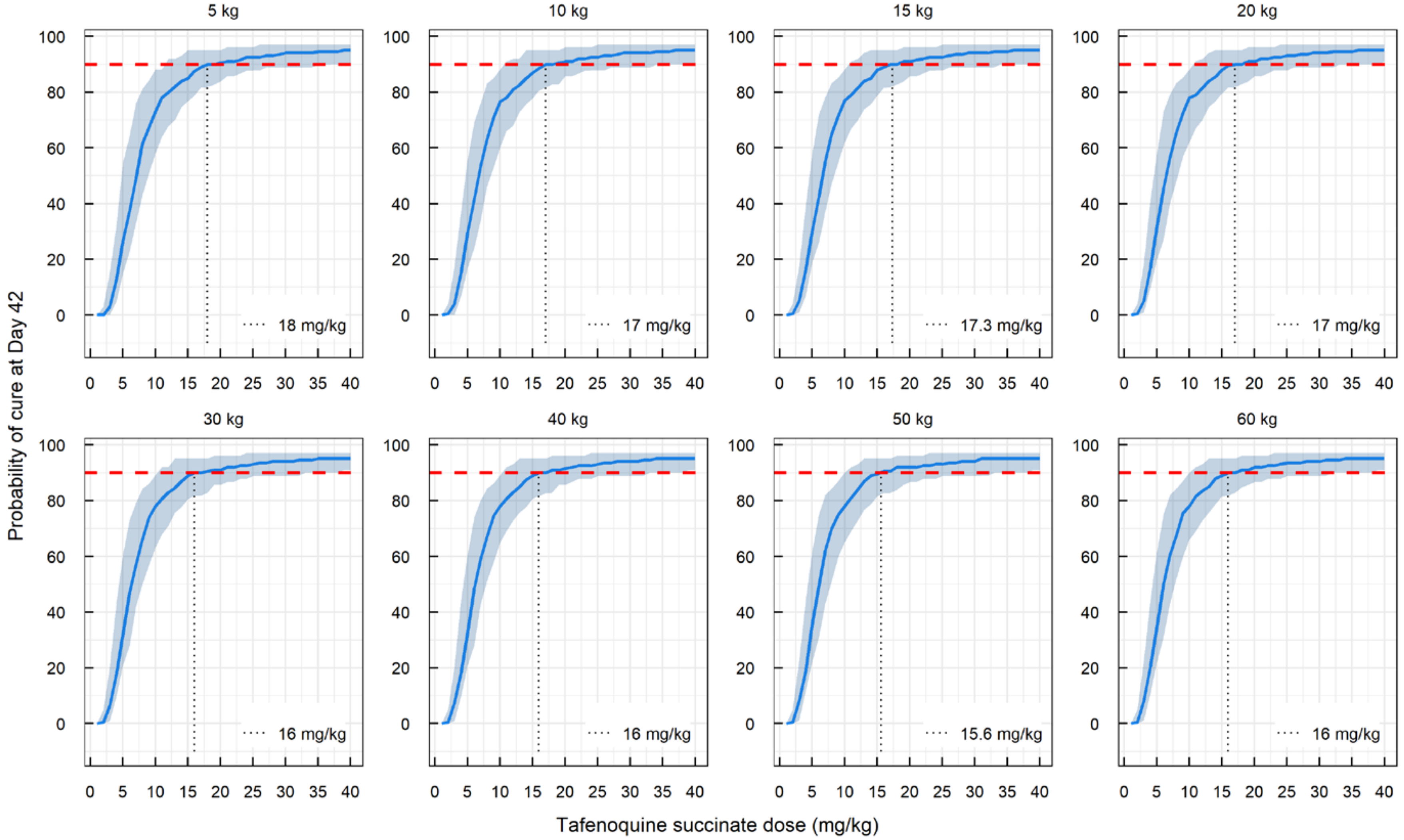

**Figure S9. Probability of adequate parasitological response within 42 days ($APR_{42}$) for different single doses of tafenoquine.**
The blue solid line represents the median $APR_{42}$; the dark blue shaded area represents the 5th-95th percentile; the red dashed line represents $APR_{42}$ at 90% probability. For each body weight, simulations were performed in 20 trials of 100 hypothetical patients each. $APR_{42}$= adequate parasitological response at Day 42.

**Table S5. Incidence of adverse events related to tafenoquine**

| Adverse event | Tafenoquine 200 mg (N=3) n (%) E | Tafenoquine 300 mg (N=4) n (%) E | Tafenoquine 400 mg (N=2) n (%) E | Tafenoquine 600 mg (N=3) n (%) E | Total (N=12) n (%) E |
|---|---|---|---|---|---|
| | **Number of participants with at least one AE (%); total number of AEs** | | | | |
| Blood Methemoglobin Present | 3 (100.0) 5 | 2 (50.0) 3 | 2 (100.0) 2 | 3 (100.0) 4 | 10 (83.3) 14 |
| Headache | 2 (66.7) 2 | 4 (100.0) 5 | 1 (50.0) 1 | 1 (33.3) 1 | 8 (66.7) 9 |
| Nausea | 0 (00.0) 0 | 2 (50.0) 2 | 1 (50.0) 1 | 2 (66.7) 2 | 5 (41.7) 5 |
| Haemoglobin Decreased | 1 (33.3) 1 | 1 (25.0) 1 | 1 (50.0) 1 | 0 (00.0) 0 | 3 (25.0) 3 |
| Abdominal Discomfort | 1 (33.3) 1 | 0 (00.0) 0 | 0 (00.0) 0 | 1 (33.3) 1 | 2 (16.7) 2 |
| Vomiting | 0 (00.0) 0 | 1 (25.0) 1 | 0 (00.0) 0 | 1 (33.3) 1 | 2 (16.7) 2 |
| Dysgeusia | 0 (00.0) 0 | 1 (25.0) 2 | 0 (00.0) 0 | 0 (00.0) 0 | 1 (8.3) 2 |
| Fatigue | 0 (00.0) 0 | 1 (25.0) 1 | 0 (00.0) 0 | 0 (00.0) 0 | 1 (8.3) 1 |
| Diarrhoea | 0 (00.0) 0 | 0 (00.0) 0 | 0 (00.0) 0 | 1 (33.3) 1 | 1 (8.3) 1 |

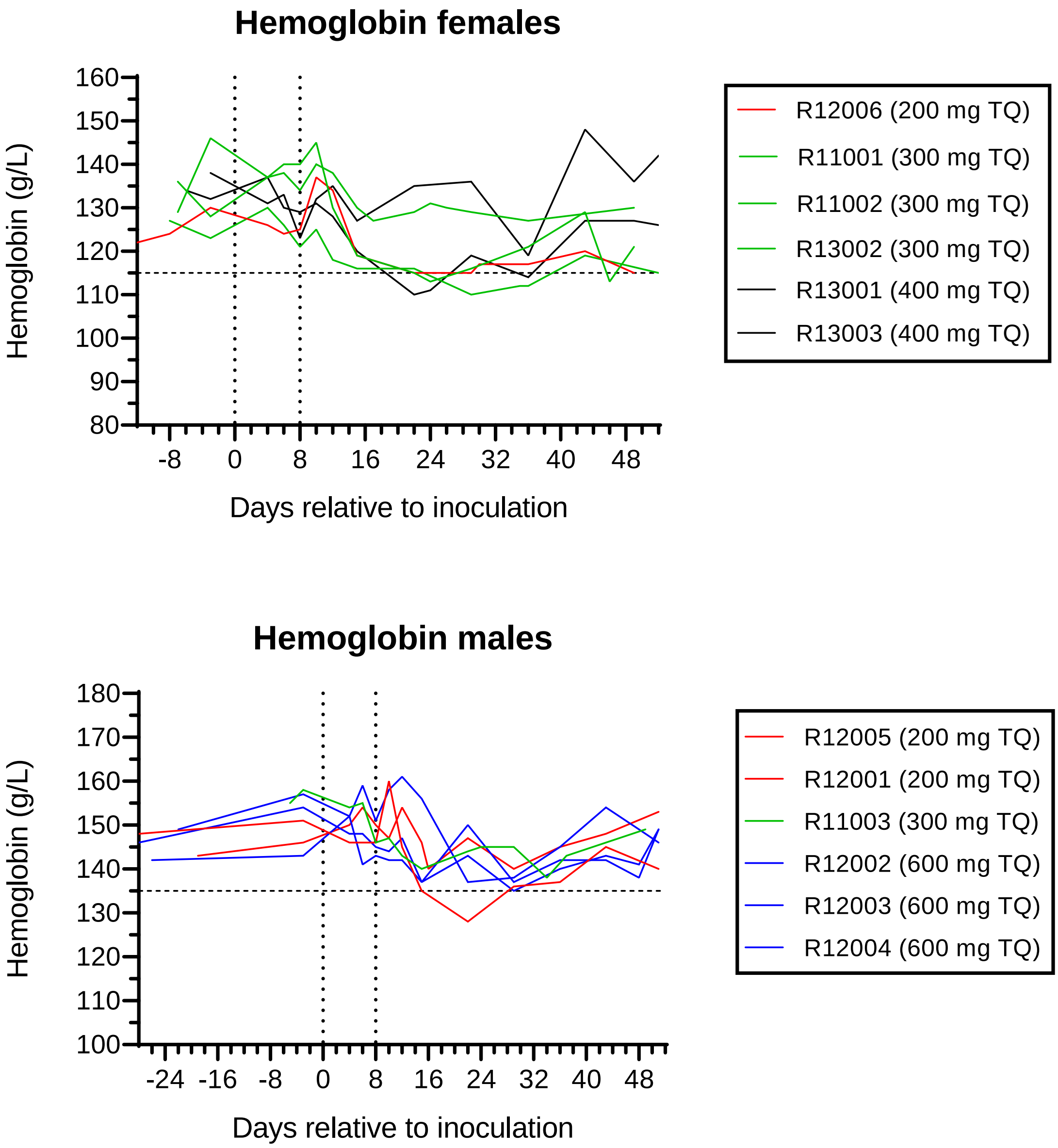

**Figure S10. Individual participant hemoglobin-time profiles**

Participants were inoculated with *P. falciparum*-infected erythrocytes on day 0 (first vertical dotted line) and were administered a single oral dose of tafenoquine on day 8 (second vertical dotted line). The horizontal dashed line represents the lower limit of the normal range for haemoglobin. TQ: tafenoquine.

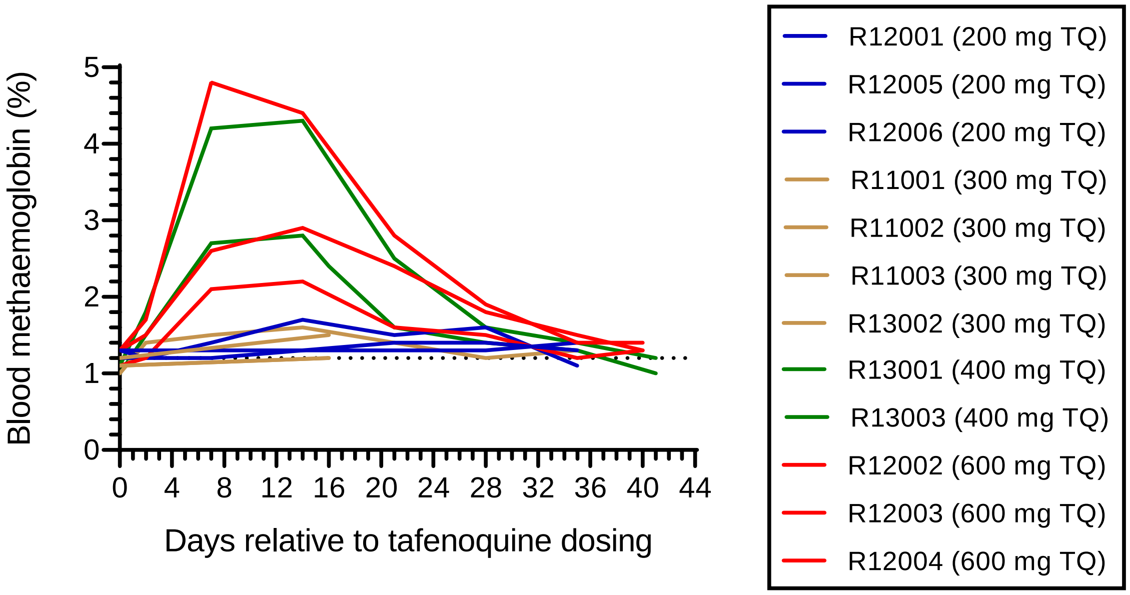

**Figure S11. Individual participant methemoglobin-time profiles**

The horizontal dotted line indicates the upper limit of the normal range of blood methemoglobin (1.2%). Blood methaemoglobin was only tested at a single time point after dosing (16 days post-dose) for participants R11001, R11002 and R11003. TQ: tafenoquine.

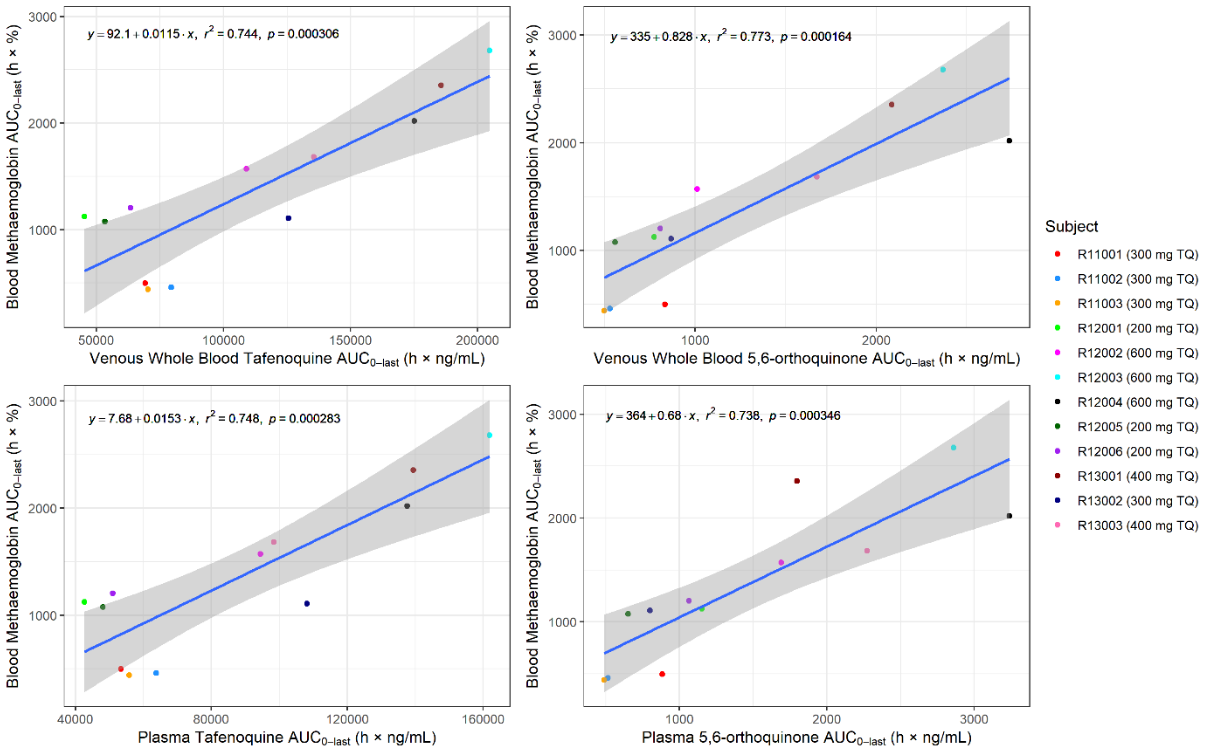

**Figure S12. Correlation between blood methemoglobin exposure and tafenoquine and 5,6-orthoquinone exposure in whole blood and plasma.**

Coloured dots represent the observed data for individual participants, blue lines represent a linear regression model, and the shaded area represents the 95% confidence interval. $AUC_{0\text{-last}}$: area under the concentration-time curve from time 0 (dosing) to the last sampling time at which the concentration is at or above the lower limit of quantification; TQ: tafenoquine.

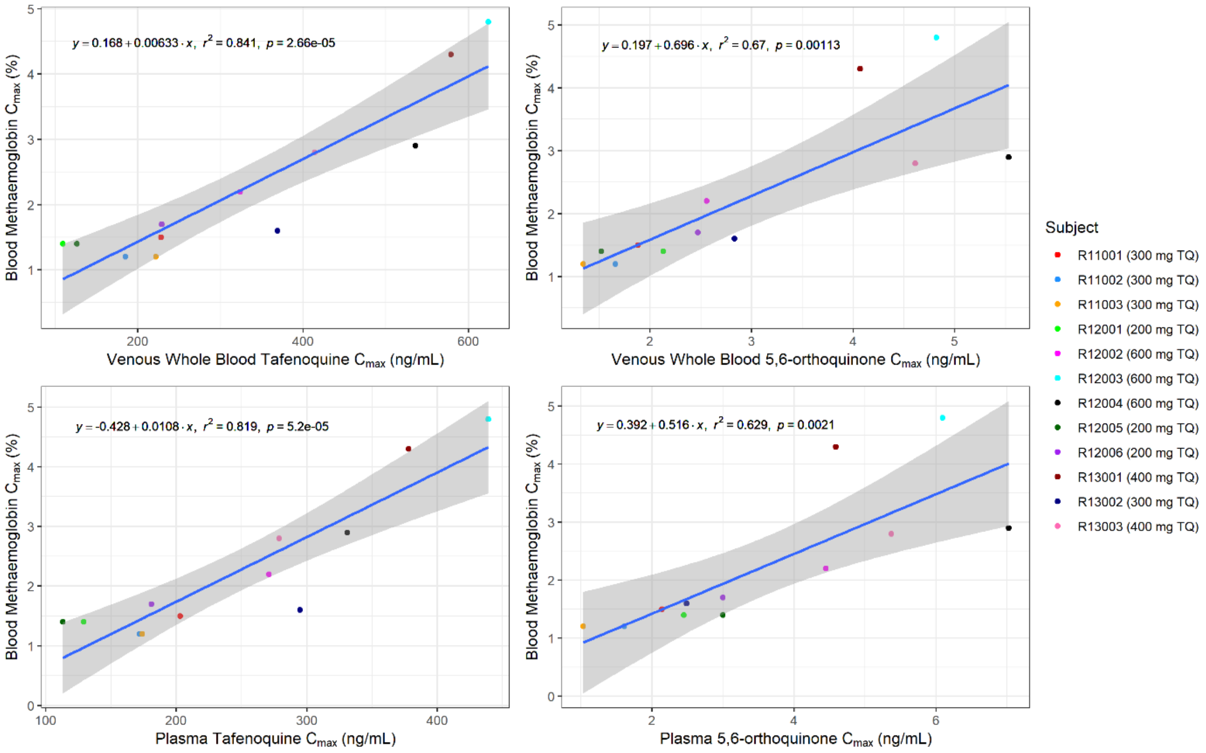

**Figure S13. Correlation between maximum blood methemoglobin concentration and maximum tafenoquine and 5,6-orthoquinone concentrations in whole blood and plasma.**

Coloured dots represent the observed data for individual participants, blue lines represent a linear regression model, and the shaded area represents the 95% confidence interval. $C_{max}$: Maximum concentration; TQ: tafenoquine.

**Text S2. Participant eligibility criteria**

### Inclusion Criteria

Volunteers who met all of the following criteria were eligible for inclusion in the study:

**Demography:**

1. Male or female (non-pregnant, non-lactating) aged 18 to 55 years inclusive who was contactable and available for the duration of the study and up to two weeks following the end of study visit.

2. Total body weight greater than or equal to 50 kg, and a body mass index (BMI) within the range of 18 to 32 kg/m$^2$ (inclusive). BMI is an estimate of body weight adjusted for height. It is calculated by dividing the weight in kilograms by the square of the height in metres.

**Health status:**

3. Certified as healthy by a comprehensive clinical assessment (detailed medical history, complete physical examination and special investigations).
4. At least normal G6PD enzyme activity levels as defined by the parameters of the specific quantitative G6PD test employed at Screening.
5. Vital signs at screening and pre-inoculation (measured after five minutes in the supine position):
    - Systolic blood pressure (SBP) 90-140 mmHg,
    - Diastolic blood pressure (DBP) 40-90 mmHg,
    - Heart rate (HR) 40-100 bpm.
6. Electrocardiograph (ECG) ranges at screening and pre-inoculation: QTcF ≤450 ms for males, QTcF ≤470 ms for females, and PR interval ≤210 ms.

7. Female volunteers of childbearing potential who had, or may have had, male sexual partner(s) during the course of the study were to use an insertable (implant and IUD, injectable, transdermal or combination oral contraceptive approved by the Australian Therapeutic Goods Administration (TGA) combined with a barrier contraceptive from the day of informed consent through to 90 days after the last dose of tafenoquine. Abstinent females were to agree to start a double method if they started a sexual relationship with a male during the study. Adequate contraception did not apply to volunteers of childbearing potential with same sex partners (abstinence from penile-vaginal intercourse), when this was their preferred and usual lifestyle. Females must not have been planning *in vitro* fertilisation within the required contraception period.
   Women of non-childbearing potential who did not require contraception during the study were defined as: surgically sterile (tubal ligation was not considered surgically sterile), post-menopausal (spontaneous amenorrhoea for ≥12 months, or spontaneous amenorrhoea for 6-12 months and follicle-stimulating hormone (FSH) ≥40 IU/mL; either was to be together with the absence of oral contraceptive use for >12 months).

   Male volunteers who had, or may have had, female sexual partner(s) during the course of the study were to agree to use a double method of contraception including condom plus diaphragm, or condom plus insertable device, or condom plus stable oral/transdermal/injectable hormonal contraceptive by the female partner, from the time of informed consent through to 90 days after the last dose of tafenoquine. Abstinent males were to start a double method if they started a sexual relationship with a female during the study, and through to 90 days after the last dose of tafenoquine. Male volunteers with female partners that were surgically sterile, or males who had undergone sterilisation and had testing to confirm the success of the sterilisation may have also been included.

**Regulations:**

8. Completion of the written informed consent process prior to undertaking any study-related procedure.

9. Must have been willing and able to communicate and participate in the whole study.
10. Agreement to adhere to the lifestyle considerations specified in the study protocol throughout the study duration.

### Exclusion Criteria

Volunteers who met any of the following criteria were not eligible for inclusion in the study:

**Medical history and clinical status:**

1. Volunteer who lives alone (at any stage from inoculation day until the end of the study). Volunteers who lived alone may have been included on a case-by-case basis, following discussion with the Principal Investigator and with approval of the Medical Monitor. Volunteers who lived alone must have identified and provided contact details of a support person who was aware of the volunteer's participation in the study and was available to provide assistance if required (for example with contacting the volunteer in the event that study staff were unable to, or with transporting the volunteer to and from the study site if required).
2. Any history of malaria or participation in a previous malaria challenge study or malaria vaccine study.
3. Must not have travelled to or lived (>2 weeks) in a malaria-endemic region during the past 12 months or planned travel to a malaria-endemic region during the course of the study. Must not have lived for >1 year in a malaria-endemic region in the past 10 years. Must not have ever lived in a malaria-endemic region for more than 10 years inclusive.
4. Had evidence of increased cardiovascular disease risk (defined as >10%, 5-year risk for those greater than 35 years of age, as determined by the Australian Absolute Cardiovascular Disease Risk Calculator (http://www.cvdcheck.org.au/)). Risk factors include sex, age, systolic blood pressure (mm/Hg), smoking status, total and HDL cholesterol (mmol/L), and reported diabetes status.
5. History of splenectomy.
6. Volunteer was unwilling to defer blood donations to the Blood Service for at least six months after the end of study visit.
7. Volunteer who had ever received a blood transfusion.
8. Any recent (<6 weeks) or current systemic therapy with an antibiotic or drug with potential antimalarial activity (e.g. chloroquine, piperaquine phosphate, benzodiazepine, flunarizine, fluoxetine, tetracycline, azithromycin, clindamycin, doxycycline etc.).
9. Known hypersensitivity to artesunate or any of its excipients, artemether or other artemisinin derivatives, proguanil/atovaquone, primaquine, or 4-aminoquinolines.
10. Hematology, clinical chemistry or urinalysis results at screening or at the day -1 to day -3 eligibility visit that were outside of Sponsor-approved clinically acceptable laboratory ranges, and were considered clinically significant by the Investigator.
11. Participation in any investigational product study within the 12 weeks preceding inoculation.
12. Symptomatic postural hypotension at screening (confirmed on two consecutive readings), irrespective of the decrease in blood pressure, or asymptomatic postural hypotension defined as a decrease in systolic blood pressure ≥20 mmHg within 2-3 minutes when changing from supine to standing position.
13. History or presence of diagnosed (by an allergist/immunologist) or treated (by a physician) food or known drug allergies (including but not limited to allergy to any of the antimalarial rescue treatments), or any history of anaphylaxis or other severe allergic reactions including face, mouth, or throat swelling or any difficulty breathing. Volunteers with seasonal allergies/hay fever or allergy to animals or house dust mite that were untreated and asymptomatic at the time of dosing could be enrolled in the study.
14. History of convulsion (including intravenous drug or vaccine-induced episodes). A medical history of a single febrile convulsion during childhood was not an exclusion criterion.
15. Presence of current or suspected serious chronic diseases such as cardiac or autoimmune disease (HIV or other immuno-deficiencies), insulin-dependent and non-insulin dependent diabetes, progressive neurological disease, severe malnutrition, acute or progressive hepatic

disease, acute or progressive renal disease, porphyria, psoriasis, rheumatoid arthritis, asthma (excluding childhood asthma, or mild asthma with preventative asthma medication required less than monthly), epilepsy, or obsessive-compulsive disorder.

16. History of malignancy of any organ system (other than localised basal cell carcinoma of the skin or *in situ* cervical cancer), treated or untreated, within five years of screening, regardless of whether there was evidence of local recurrence or metastases.
17. History of schizophrenia, bi-polar disease, psychoses, disorders requiring lithium, attempted or planned suicide, or any other severe (disabling) chronic psychiatric diagnosis.
18. Volunteers who had received psychiatric medications within one year prior to enrolment for a psychiatric condition, or who had been hospitalised within five years prior to enrolment for either a psychiatric illness or due to danger to self or others.
19. History of an episode of minor depression that required at least six months of pharmacological therapy and/or psychotherapy within the last five years; or any episode of major depression. The Beck Depression Inventory was used as an objective tool for the assessment of depression at screening. In addition to the conditions listed above, volunteers with a score of 20 or more on the Beck Depression Inventory and/or a response of 1, 2 or 3 for item 9 of this inventory (related to suicidal ideation) were not eligible for participation. These volunteers were to be referred to a general practitioner or medical specialist as appropriate. Volunteers with a Beck score of 17 to 19 may have been enrolled at the discretion of the Investigator if they did not have a history of the psychiatric conditions mentioned in this criterion and their mental state was not considered to pose additional risk to the health of the volunteer or to the execution of the study and interpretation of the data gathered.
20. History of recurrent headache (e.g. tension-type, cluster or migraine) with a frequency of ≥2 episodes per month on average and severe enough to require medical therapy, during the 5 years preceding screening.
21. Presence of clinically significant infectious disease or fever (e.g. sublingual temperature ≥38.5°C) within the five days prior to inoculation.
22. Evidence of acute illness within the four weeks prior to screening that the investigator deemed may have compromised volunteer safety.
23. Significant inter-current disease of any type, in particular liver, renal, cardiac, pulmonary, neurologic, rheumatologic, or autoimmune disease by history, physical examination, and/or laboratory studies including urinalysis.
24. Volunteer had a clinically significant disease or any condition or disease that might affect drug absorption, distribution or excretion (e.g. gastrectomy, diarrhoea).
25. Blood donation of any volume or participation in any research study involving blood sampling (more than 450 mL/unit of blood) within one month before tafenoquine administration, or blood donation to Australian Red Cross Blood Service (Blood Service) or other blood bank during the eight weeks prior to inoculation.
26. Medical requirement for intravenous immunoglobulin or blood transfusions.
27. History or presence of alcohol abuse (alcohol consumption more than 40 g/4 units/4 standard drinks per day), or drug habituation, or any prior intravenous usage of an illicit substance.
28. Female volunteer who was breastfeeding.
29. Any vaccination within the last 28 days.
30. Any corticosteroids, anti-inflammatory drugs (excluding commonly used over-the-counter anti-inflammatory drugs such as ibuprofen, acetylsalicylic acid, diclofenac), immunomodulators or anticoagulants within the past three months. Any volunteer currently receiving or having previously received immunosuppressive therapy (including systemic steroids, adrenocorticotrophic hormone or inhaled steroids) at a dose or duration potentially associated with hypothalamic-pituitary-adrenal axis suppression within the past year.
31. Use of prescription drugs (excluding contraceptives) or non-prescription drugs or herbal supplements (such as St John's Wort), within 14 days or five half-lives (whichever was longer) prior to inoculation. Limited use of other non-prescription medications or dietary supplements, not believed to affect volunteer safety or the overall results of the study, may have been permitted on a case-by-case basis following approval by the sponsor in consultation with the

investigator. Volunteers were requested to refrain from taking non-approved concomitant medications from recruitment until the conclusion of the study.

32. Cardiac/QT risk:
    - Family history of sudden death or of congenital prolongation of the QTc interval or known congenital prolongation of the QTc interval or any clinical condition known to prolong the QTc interval.
    - History of symptomatic cardiac arrhythmias or with clinically relevant bradycardia.
    - Electrolyte disturbances, particularly hypokalaemia, hypocalcaemia, or hypomagnesaemia.
    - ECG abnormalities in the standard 12-lead ECG (at screening and prior to inoculation) which in the opinion of the Investigator was clinically relevant or would interfere with the ECG analyses.
33. History of retinal abnormalities, diseases of the retina or macula of the eye, visual field defects, and hearing disorders like reduced hearing and tinnitus.

**General conditions:**

34. Any volunteer who, in the judgment of the investigator, was likely to be non-compliant during the study, or was unable to cooperate because of a language problem or poor mental development.
35. Any volunteer in the exclusion period of a previous study according to applicable regulations.
36. Any volunteer who was the investigator, research assistant, pharmacist, study coordinator, or other staff thereof, directly involved in conducting the study.
37. Any volunteer without good peripheral venous access.

**Biological status:**

38. Positive result on any of the following tests: hepatitis B surface antigen (HBs Ag), anti-hepatitis B core antibodies (anti-HBc Ab), anti-hepatitis C virus (anti-HCV) antibodies, anti-human immunodeficiency virus 1 and 2 antibodies (anti-HIV1 and anti-HIV2 Ab).
39. Positive urine drug test. Any drug in the urine drug screen unless there was an explanation acceptable to the investigator (e.g., the volunteer had stated in advance that they had consumed a prescription or over-the-counter product which contained the detected drug) and/or the volunteer had a negative urine drug screen on retest by the pathology laboratory. Any volunteer testing positive for acetaminophen (paracetamol) at screening and/or inoculation day may still have been eligible for study participation, at the investigator’s discretion.
40. Positive alcohol breath test.
